# Efficacy and Safety of Habitual Consumption of a Food Supplement Containing Miraculin in Malnourished Cancer Patients: the CLINMIR Pilot Study

**DOI:** 10.1101/2024.05.15.24307453

**Authors:** Bricia López-Plaza, Ana Isabel Álvarez-Mercado, Lucía Arcos-Castellanos, Julio Plaza-Diaz, Francisco Javier Ruiz-Ojeda, Marco Brandimonte-Hernández, Jaime Feliú-Batlle, Thomas Hummel, Ángel Gil, Samara Palma Milla

**Author notes:** These authors contributed equally to this work (Ángel Gil, Samara Palma Milla). Correspondence; (B.L.-P).

## Abstract

Taste disorders (TDs) are common among systemically treated cancer patients and negatively impact their nutritional status and quality of life. A food supplement containing the natural taste-modifying protein miraculin (DMB^®^) has emerged as a possible alternative treatment for TDs. The present study aimed to evaluate the efficacy and safety of habitual DMB consumption in malnourished cancer patients undergoing active treatment. An exploratory clinical trial was carried out in which 31 cancer patients were randomized into three arms [standard dose of DMB (150 mg DMB/tablet), high dose of DMB (300 mg DMB/tablet) or placebo (300 mg freeze-dried strawberry)] for three months. Patients consumed an intervention DMB tablet or placebo before each main meal. Throughout the five main visits, electrochemical taste perception, nutritional status, dietary intake, quality of life and the fatty acid profile of erythrocytes were evaluated. Patients consuming a standard dose of DMB exhibited improved taste acuity over time (% change right/left side: ‒52.8 ± 38.5 / ‒58.7 ± 69.2%) and salty taste perception (2.29 ± 1.25 *vs.* high dose: 2.17 ± 1.84 *vs.* placebo: 1.57 ± 1.51 points, p < 0.05). They also had higher energy intake (p = 0.075) and covered better energy expenditure (107 ± 19%). The quality of life evaluated by symptom scales improved in patients receiving the standard dose of DMB (constipation, p = 0.048). The levels of arachidonic (13.1 ± 1.8; 14.0 ± 2.8, 12.0 ± 2.0%; p = 0.004) and docosahexaenoic (4.4 ± 1.7; 4.1 ± 1.0; 3.9 ± 1.6%; p = 0.014) acids in erythrocytes increased over time after DMB intake. The standard dose of DMB increased fat‒free mass *vs*. placebo (47.4 ± 9.3 vs. 44.1 ± 4.7 kg, p = 0.007). Importantly, habitual patients with DMB did not experience any adverse events, and metabolic parameters remained stable and within normal ranges. In conclusion, habitual consumption of a standard 150 mg dose of DMB improves electrochemical food perception, nutritional status (energy intake, fat quantity and quality, fat-free mass) and quality of life in malnourished cancer patients receiving antineoplastic treatment. Additionally, DMB consumption appears to be safe, with no changes in major biochemical parameters associated with health status. The clinical trial was registered at http://clinicaltrials.gov (NCT05486260).

## 1. Introduction

Taste disorders (TDs) are frequent adverse events during antineoplastic treatments in cancer patients [1–4]. However, limited attention has been given to these disorders. The effects of TDs are related to the cytotoxic effects of chemotherapy on the differentiation and proliferation of cells in the taste bud [5] or to chemosensory dysfunction that can cause neurological damage by acting directly on taste receptors or synaptic uncoupling during radiotherapy [6]. Stem cell therapy [7] and anticancer-targeted drugs [8,9] have also been shown to induce taste alterations. However, chemotherapy-related TDs are more frequent. Chemotherapy-induced TDs are highly variable and range between 17% and 86% [10]. The presence of TDs can occur as acute side effects after chemotherapy [11] increasing according to the number of cycles received. Although these symptoms generally improve once treatment is completed [12], they may also persist for a long period after treatment is completed [13]. One of the most prevalent TDs is dysgeusia, which occurs between 56% and 76% of patients receiving antineoplastic treatment [14]. Dysgeusia is a qualitative gustatory disturbance defined as impaired or altered sense taste perception or persistent taste sensation without stimulation [15]. Generally, patients described unpleasant tastes or distortions of taste sensation [16].

Patients commonly present anorexia due to antineoplastic treatment but also due to dysgeusia. Indeed, patients attribute difficulties maintaining adequate food intake to altered taste during treatment [17]. TDs reduce appetite and energy intake, which produce changes in food preferences [18] that determine weight loss and changes in body composition [19] and increase malnutrition risk in cancer patients [20].

The prevalence of malnutrition in cancer patients varies between 40% and 80% [21]. This condition determines the outcome in cancer patients [22] since its presence is associated with treatment–induced toxicity, an increase in the postoperative risk of complications [23], poor prognosis, overall survival reduction [24] and increased mortality. In this sense, TDs can increase malnutrition risk by a factor of 3.36 [19]. TDs can also have a significant impact on cancer patients’ quality of life by reducing food enjoyment [25,26] and developing food aversions that reduce food intake [27] and increase the risk of malnutrition [28,29].

Therefore, it is not surprising that different strategies have been developed to prevent or ameliorate TDs [30–34]. Commonly known as the miracle berry, the *Synsepalum dulcificum* (Daniell) fruit has attracted increased attention due to its ability to transform sour taste perception into sweet taste [35]. This quality is due to the presence of miraculin, a glycoprotein that acts as a selective agonist at acidic pH or antagonist at neutral pH, of sweet taste receptors [36]. This characteristic allows miraculin to change the food flavor depending on the pH of the food consumed making meals more palatable. Miraculin provides a high sweetness intensity that persists for approximately 30 minutes after consumption [37]; thus, its consumption could improve the overall taste perception in cancer patients undergoing antineoplastic treatment and those with TDs [38], improving food intake and, consequently, their nutritional and health status.

Two studies have evaluated the consumption of miracle fruit in cancer patients undergoing active chemotherapy treatment, and both have shown positive changes in TDs [39,40]. However, despite pointing out the direction of the effect of consuming the miracle berry on these patients, both studies used subjective methods for the assessment of TDs and used the fruits of *S. dulcificum*.

In December 2021, the European Commission authorized dried miracle berry (DMB) as a *novel food* [41]. DMB, is a freeze-dried extract of miracle berry pulp juice rich in miraculin. It was officially cataloged as the dried fruit of *S. dulcificum*, safe for use in the European Union. DMB^®^ has become available as a food supplement.

In this sense, the present study hypothesizes that DMB consumption enhances the electrochemical taste perception and improves both the nutritional status and quality of life of cancer patients positively impacting their health. Therefore, the main aim of the present clinical trial was to evaluate the efficacy and safety of habitual DMB consumption in malnourished cancer patients undergoing active treatment.

## 2. Materials and Methods

A detailed description of the CLINMIR study protocol has recently been published elsewhere [42]. Below is a summary of the clinical trial.

### 2.1 Trial design

The clinical trial protocol was approved by the Scientific Research and Ethics Committee of the Hospital University La Paz (HULP), Madrid (Spain) in version 1 in June 2022 and protocolled by the HULP Code 6164. The present protocol clinical trial has also been registered at http://clinicaltrials.gov with the number NCT05486260.

The CLINMIR study is a pilot randomized, parallel, triple-blind, and placebo-controlled clinical trial allocated in three arms according to treatment with a food supplement enriched in the protein miraculin (DMB) in malnourished cancer patients exhibiting TDs because of active chemotherapy and radiotherapy and adjusted by type of cancer. All patients were recruited from medical consultations in the Clinical and Dietary Nutrition Unit (UNC&D) and by referral from the Oncology Service of the HULP to UNC&D.

### 2.2 Participants

The main inclusion criteria were patients 18 years of age and older with cancer, active chemotherapy and/or radiotherapy, and/or immunotherapy treatment who had a weight loss ≥ 5 % in the last six months, malnutrition diagnosis assessed by Global Leadership Initiative on Malnutrition (GLIM Criteria) [43], and TDs measured by electrogustometry. Additionally, patients had to have a life expectancy greater than 3 months and be able to feed by oral intake. Patients also had an understanding of the clinical study guidelines.

The exclusion criteria included patients participating in another clinical trial, enteral or parenteral nutrition, poorly controlled diabetes mellitus (HbA1c >8%), uncontrolled hypertension or hyper/hypothyroidism, severe digestive toxicity due to treatment with chemo-radiotherapy, severe kidney or liver disease (chronic renal failure, nephrotic syndrome, cirrhosis, etc.), severe dementia, brain metastases, eating disorders, history of severe neurological or psychiatric pathology that may interfere with treatment, alcoholism or substance abuse, severe gastrointestinal diseases, and unwillingness to consume the miraculin-based food supplement.

Intolerance to miraculin was a withdrawal criterion. Any medication that did not interfere with the study formulation was allowed and registered in the Clinical Research Data.

### 2.3 Interventions

Patients who met the selection criteria were randomized to one of three arms of the clinical trial. The first arm had 150 mg of DMB equivalent to 2.8 mg of miraculin + 150 mg of freeze-dried strawberries per orodispersible tablet; the second arm had 300 mg of DMB equivalent to 5.6 mg of miraculin; and the third arm contained 300 mg of freeze-dried strawberries per orodispersible tablet as a placebo. All treatments were isocaloric (**Table 1**).

**Table 1.**
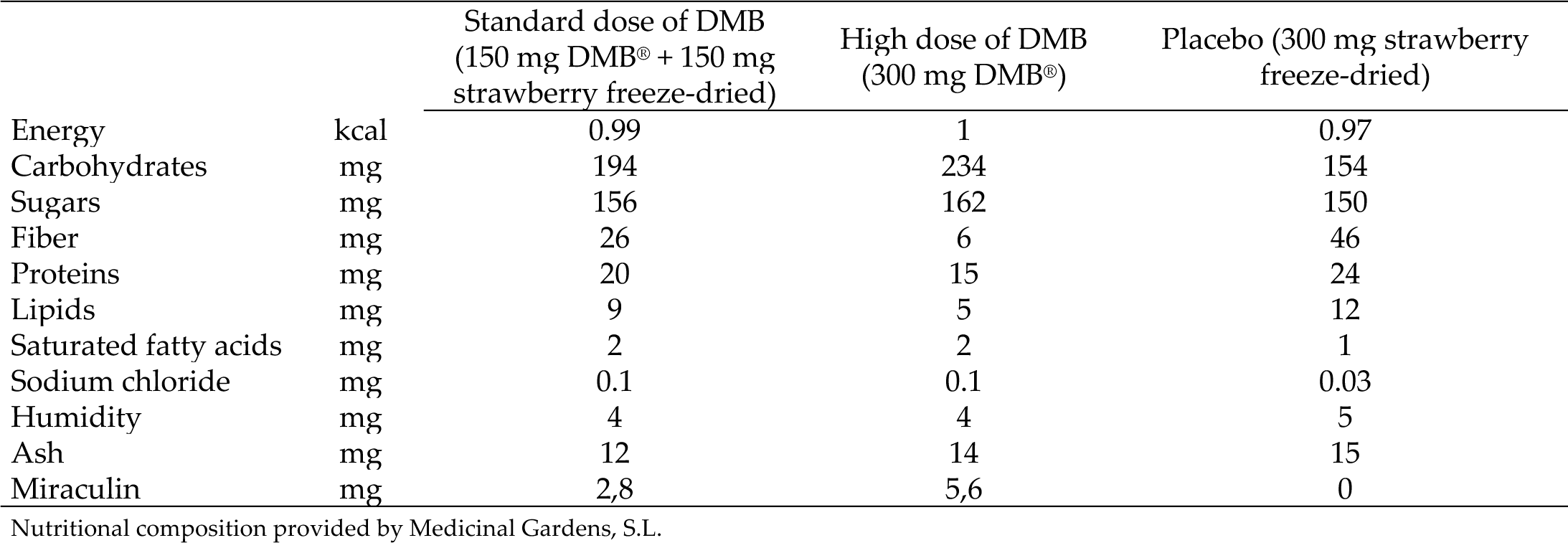
Nutritional composition of the food supplement enriched in miraculin (DMB) and placebo.

|  |  | Standard dose of DMB<br>(150 mg DMB® + 150 mg<br>strawberry freeze-dried) | High dose of DMB<br>(300 mg DMB®) | Placebo (300 mg strawberry<br>freeze-dried) |
| --- | --- | --- | --- | --- |
| Energy | kcal | 0.99 | 1 | 0.97 |
| Carbohydrates | mg | 194 | 234 | 154 |
| Sugars | mg | 156 | 162 | 150 |
| Fiber | mg | 26 | 6 | 46 |
| Proteins | mg | 20 | 15 | 24 |
| Lipids | mg | 9 | 5 | 12 |
| Saturated fatty acids | mg | 2 | 2 | 1 |
| Sodium chloride | mg | 0.1 | 0.1 | 0.03 |
| Humidity | mg | 4 | 4 | 5 |
| Ash | mg | 12 | 14 | 15 |
| Miraculin | mg | 2,8 | 5,6 | 0 |

Nutritional composition provided by Medicinal Gardens, S.L.

Those patients who voluntarily agreed to participate signed the informed consent form. Over 3 months, each patient consumed an orodispersible tablet containing DMB or placebo five minutes before each main meal (breakfast, lunch, and dinner).

The clinical trial had six face-to-face visits in two phases, one selection visit (vS) in the Selection Phase and five visits in the Experimental Phase (**Figure 1**).

**Figure 1.**
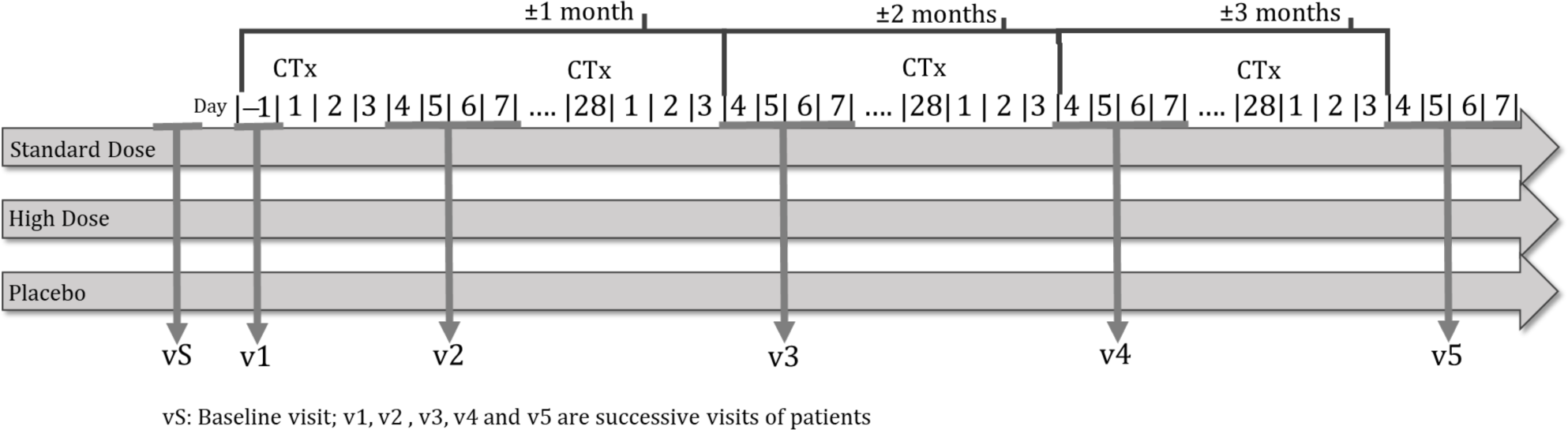
CLINMIR clinical trial outline

On the selection visit, nutritional status was assessed according to the GLIM criteria as well as electrical (electrogustometry) and chemical taste perception (taste strips). The included patient received the questionnaires to complete and hand in at visit 1 (food daily record of 3 days, one holiday, quality of life questionnaire -EORTC QLQ-C30- and International Physical Activity Questionnaire-IPAQ-as well as the blood sample extraction appointment (analysis of biochemical parameters and fatty acids from erythrocytes). Experimental phase visits were 4-7 days after their chemotherapy infusion, except visit 1 before it.

At visit 1 (v1) patients were randomized and provided with the necessary product (DMB or placebo) until their next visit (v2). Anthropometric measurements, electrical bioimpedance and the Sniffin’ Sticks Smell Test were carried out. Healthy eating and physical exercise guidelines for cancer patients were explained. As part of the next visit, the following forms were delivered: a product efficacy satisfaction questionnaire, a product consumption control daily sheet, a product consumption tolerance record sheet, and a record sheet of adverse effects. Additionally, individualized nutritional treatment was implemented. If an oral nutritional supplement was needed, a polymeric, hypercaloric, and hyperproteic formula enriched in omega-3 fatty acids was prescribed depending on their energy requirements.

Visits 2 (v2, 4−7 days after the chemotherapy session), 3 (v3, ±1 month after visit 1) and 4 (v4, ±2 months after visit 1) were similar and they were carried out 4−7 days after the chemotherapy session. During these visits, nutritional status was monitored, and anthropometric measurements and smell and taste tests (electrogustometry, taste strips tests and Sniffin’ sticks smell test) were carried out. In these visits, biochemical parameters were also measured. Completed questionnaires were collected (food daily record, quality of life questionnaire, product efficacy, product consumption control, tolerance record and adverse effects record) and behavioral reinforcement (nutritional treatment and physical activity, consumption and registration of the assigned treatment, and tolerance and adverse effects registry). Patients received the questionnaires to complete and hand in at the next visit.

Finally, during visit 5 (v5, ±3 months after v1 and 3-4 days after the patient’s chemotherapy) nutritional status was assessed and anthropometric measurements and taste and smell tests were carried out (electrogustometry, taste strips tests and Sniffin’ sticks smell test). A blood sample was extracted (biochemical parameters and fatty acids from erythrocytes) for analysis. Food daily records and quality of life questionnaire completed were collected as well as a product efficacy questionnaire, product consumption control, tolerance record and adverse effects record. Behavioral reinforcement of nutritional treatment and physical activity were carried out.

### 2.4 Outcomes

Malnourished cancer patients with TDs and consuming DMB were expected to improve their taste perception by reducing the electrical–chemical taste perception threshold from baseline (v0) and throughout the intervention. Moreover, it is expected that DMB consumption improves the chemical and olfactory perception of food. Improvements in dietary intake and nutritional and safety biochemical parameters, as well as improvements in the essential and polyunsaturated fatty acid status assessed through the fatty acid composition of erythrocytes, were expected because of a better perception of food. Tolerance and possible adverse effects were also outcomes studied since several doses were evaluated. All parameters were evaluated from baseline to the end of the intervention and evolution was measured through the different visits carried out (v1, v2, v3, v4, v5).

### 2.5 Sample size

Because the CLINMIR study was exploratory and there was a lack of previous studies using miraculin-based nutrition supplements in cancer patients, the sample size was established by the researchers. The number established was 10 patients per arm given a sample size of 30 patients. The results obtained will be able to serve to establish the sample size needed to evaluate the efficacy of the intervention product in multicenter studies.

### 2.6 Randomization and Blinding

Randomization was carried out using computer-generated random numbers in blocks of six taking into account treatment and cancer type. This sequence was generated by the Biostatistics Unit (HULP). The allocation sequence was provided in a separate document. To implement the allocation, the sequences were sequentially numbered and sealed in envelopes that were mailed to the nutritionist who enrolled and assigned participants to interventions. When the patient signed the informed consent (v1) patient’s randomization envelope was opened.

Researchers, trial patients, care providers (nutritionists, nurses, physicians), assessing outcomes, data analysts, and the promoter were blinded after assignment to interventions. Both miraculin-based food supplements and placebo had similar appearances (pink tablets). They were packaged in white opaque bottles with 30 orodispersible tablets identified by a lot number (L01, L02, L03) and a barcode for tracking. The test product in its powder form (DMB^®^) and the placebo were provided by Baïa Food (Medicinal Gardens SL) to Rioja Nature Pharma. The packaging, in the form of bottles equipped with protective technology for moisture and oxygen-sensitive products (Activ Vial^®^), was supplied by CSP Aptar Technologies. Rioja Nature Pharma was responsible for the manufacturing, labeling, identification, and supply of the final product, and maintained the blind throughout the study until the statistical analysis was completed.

### 2.7 Specific methodology

#### 2.7.1 Malnutrition criteria

Nutritional diagnosis of malnutrition was established through the GLIM criteria based on phenotypic and etiological criteria. It requires at least one phenotypic criterion and one etiologic criterion to diagnose malnutrition. Body composition by bioelectrical impedance analysis (BIA) was used to evaluate reduced muscle mass. Gastrointestinal symptoms as supportive indicators were considered to assess to evaluate reduced food assimilation and major infection. Finally, trauma or acute conditions were associated with inflammation. Malnutrition was classified as moderate or severe malnutrition [43]. Nutritional status was evaluated at all study visits.

#### 2.7.2 Anthropometric parameters

They were taken using standard techniques, following the international norms established by the WHO. Body weight was measured using a clinical digital scale (capacity 0-150 kg). The percentage of weight loss was assessed as follows: [(current weight ‒ weight 6 months ago)/weight 6 months ago] * 100. Height was measured with a height meter with an accuracy of 1 mm (range, 80-200 cm). Body mass index (BMI) was determined using weight (kg)/height (m)^2^. Anthropometric parameters were measured at the main visits (v1, v3, v4 & v5).

#### 2.7.3 Daily food record

Diet was collected in three different days’ daily food records, one of which had to be a holiday. Patients were instructed to record the weight of the food consumed or, if this was not possible, to record household measurements (spoonfuls, cups, etc.). All records were thoroughly reviewed by a nutritionist in the presence of the patient to ensure that the information collected was complete. Foods, drinks, dietary supplements, and preparations consumed were transformed into energy and nutrients using DIAL software (Alce Ingeniería, Madrid, Spain). Results were compared with the recommended intakes of the Spanish population.

#### 2.7.4 Electrogustometry

The threshold for an electric-induced taste stimulus (taste acuity) was measured using an electrogustometer (SI-03 Model, Sensonics International, New Jersey, USA). Patients were instructed not to eat or drink for an hour before electrogustometry. Monopolar electrode applied electric stimulus. The electrogustometer produces low-amplitude stimuli of a predetermined duration (0.5 seconds). The methodology used was that recommended by the manufacturer. The electric threshold scores were measured in the area of the fungiform papillae on both sides of the tongue. To detect thresholds, a two-down and one-up forced-choice single staircase procedure and a stimulus-response staircase were used. Threshold differences between the left and right sides greater than 7 dB were considered abnormal [44].

#### 2.7.5 Taste strips test

A validated method to measure chemical taste perception [30,134] this tool is based on the chemical perception of taste through taste-impregnated filter paper strips (Burghart Messtechnik GmbH, Germany). Four different taste strips (sweet, sour, salty, and bitter) were measured at four different concentrations each. For the assessment of whole-mouth gustatory function, strips were placed on the tongue and savored with the closed mouth for 10 seconds. Once the strip was removed, the participants had to identify the taste within a forced choice procedure. A maximum score of 16 points (4 concentrations of each of the 4 basic taste qualities) was obtained. Hypogeusia was considered when a score below nine was obtained regardless of age.

#### 2.7.6 Sniffin’ Sticks Smell test

Smell perception was measured based on odor-containing felt-tip pens (“Sniffin’ sticks” Burghart Messtechnik GmbH, Germany). Consuming food, drinks, or cigarettes 15 minutes before testing was not allowed. A total of 16 odor pens were presented to be identified. For each pen, a flash card with 4 choices was provided (e.g., pineapple, orange, blackberry, strawberry). Each uncovered odor pen was held 2 cm in front of the nostrils for 3‒4 seconds. Based on the multiple forced choice paradigm patients had to choose the best match with their olfactory perception. The score sums all correct answers and was used to differentiate between normosmia and hyposmia depending on the age of the patient.

### 2.8 Quality of life

This was evaluated using the EORTC QLQ-C30 questionnaire for cancer patients validated in Spanish [45]. The questionnaire is formed by 5 functional scales (daily activities and physical, emotional, cognitive, and social functioning), 3 symptomatic scales (fatigue, pain and nausea, and vomiting), 1 overall health scale, and 6 questions about dyspnea, insomnia, anorexia, constipation, diarrhea, and economic impact. All questions are about the previous week and are scored with 1 to 4 points. The last two questions have a score from 1 to 7, with 1 being terrible and 7 being excellent.

Scores obtained are standardized from 0 to 100 points to determine the disease impact on each scale. High scores on the global health status and functional scales indicate a better quality of life, while low scores on the symptoms scale indicate a decrease in quality of life.

### 2.9 Tolerance and adverse events

Gastrointestinal disorders such as abdominal distension, abdominal pain, nausea, regurgitation or gastroesophageal reflux, vomiting, constipation, diarrhea, and flatulence were defined and recorded based on the Common Terminology Criteria for Adverse Events (CTCAE) from the National Cancer Institute [46]. These adverse events were classified as Grade 0 (not described), Grade 1 (mild), Grade 2 (moderate), Grade 3 (severe), Grade 4 (mortality risk), and Grade 5 (death associated with an event). Additionally, the patients were asked if they could be related to product consumption.

### 2.10 Fatty acid profile of erythrocytes

The separation and quantification of fatty acids from erythrocyte lipids have been reported in previous works [47]. Briefly, erythrocyte lipid extraction and fatty acid methylation were performed as described by Lepage & Roy (1988) [48]. Fatty acid methyl esters (FAME) were identified and quantified by comparing their retention times by gas chromatography-mass spectrometry (GC-MS). This analysis was performed by injecting 1µl into a Bruker (Bremen, Germany) model 456-GC high-resolution gas chromatograph coupled to a Bruker model EVOQ TQ triple quadrupole mass spectrometer as follows:

GC conditions

a. ZB-FAME capillary column (30m x 0.25mm ID x 0.20um film).
b. Split mode injector (100:1)
c. Injector temperature: 250°C
d. Transfer line temperature: 240°C
e. Carrier gas: He (1 ml/min)
f. Temperature ramp: 100°C (2 min) up to 210°C (5 min) at 4°/min.

MS conditions:

a. Temperature of the source: 240°C
b. Full scan from 45 Da to 450 Da
c. Electron impact ionization (EI+) at 70eVFood daily record

### 2.11 Biochemical parameters

Biochemical analyses were carried out in the Biochemistry Laboratory of the Hospital La Paz, an ISO-certified laboratory, at each visit (v1, v3, v4, v5) using an Olympus AU5400 Automated Chemistry Analyzer (Olympus Corporation, Izasa, CA, USA).

### 2.12 Miraculin-based food supplement taste perception

A visual analog scale (VAS) was designed by the researchers to obtain information about the miraculin-based food supplement’s taste perception efficacy. The questionnaire included 5 questions using 10 cm scales, where 0 means not at all or very bad and 10 means very good or very effective. The questions included were as follows: Do you notice a food taste change after consuming the product? Does food taste better to you? Does it allow you to eat more food? What is your opinion of the product? Are you satisfied with the effectiveness of the product? Does the administration of the product seem adequate to you?

### 2.13 Statistical methods

Data analysis was carried out by the intention to treat. Quantitative data are presented as the means ± standard deviations (SD), and percentages. Data type distribution was determined using Shapiro‒Wilks tests. Levene’s test was used to evaluate the homogeneity of variances. Parametric or nonparametric tests were performed depending on the data distribution. General linear mixed models (GLM) of covariance (ANCOVA) were used to evaluate differences between means for treatment, time, and treatment x time using as covariates the baseline data. The analysis of the qualitative variables and percentages was carried out through χ2 or Fisher’s F analysis.

Double-sided tests were applied when needed, and a p-value < 0.05 was considered statistically significant. Data were analyzed using R Project for Statistical Computing (https://www.r-project.org/).

## 3. Results

The recruitment period was extended from November 2022 to May 2023. A total of 62 patients were evaluated for eligibility. Of them, 31 oncologic patients met the selection criteria and were randomized into the three intervention groups, adjusted by the type of cancer (**Figure 2**). During follow-up, extended from November 2022 to August 2023, there were 10 dropouts, most of them due to the taste distortion of non-sweet acidic foods (n = 6) and because the prescription derived from the intervention added difficulty to their, already complex, antineoplastic treatment (n = 2). Additionally, there were 2 *exitus letalis* in the placebo group. There was a 32 % dropout and only 21 cancer patients completed the clinical trial; however, all variables were evaluated by intention to treat.

**Figure 2.**
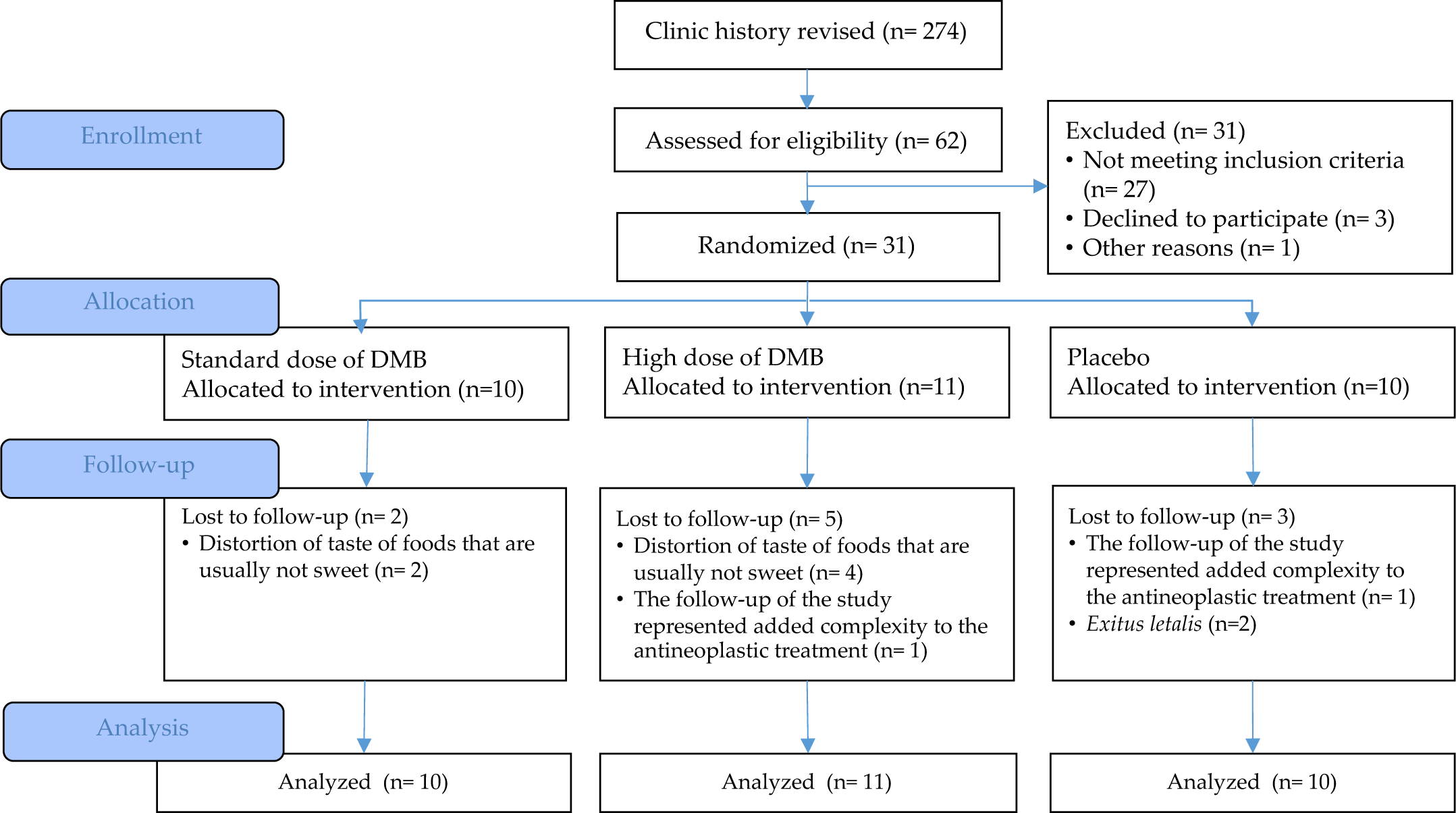
CONSORT flow diagram.

### 3.1 General characteristics of the population

The sample was made up of 58.1 % women and 41.9 % men with a mean age of 60.0 ± 10.9 years old, all of them undergoing active treatment with at least chemotherapy, and TDs were measured by electrogustometry (**Table 2**).

**Table 2.** Baseline characteristics of the population.

|  |  | Standard dose of DMB | High dose of DMB | Placebo | p-value |
| --- | --- | --- | --- | --- | --- |
| Sex | Female (%) | 70 | 45.5 | 60 | 0.517 |
|  | Male (%) | 30 | 54.5 | 40 |  |
| Age | years | 59.9 ± 15.1 | 58.9 ± 4.9 | 61.3 ± 11.2 | 0.891 |
| Weight | kg | 61.4 ± 11.1 | 62.0 ± 14.1 | 62.6 ± 10.7 | 0.941 |
| Weight lost in last 6 mo. | % | 7.5 ± 6.0 | 8.7 ± 7.1 | 7.2 ± 8.0 | 0.868 |
| BMI | kg/m<sup>2</sup> | 21.9 ± 3.6 | 22.0 ± 3.3 | 22.9 ± 3.4 | 0.737 |
| Type of cancer |  |  |  |  |  |
|  | Head and neck | 0 | 9.1 | 0 | 0.895 |
|  | Colorectal | 30 | 27.3 | 20 |  |
|  | Esophagus | 10 | 0 | 10 |  |
|  | Stomach | 0 | 9.1 | 10 |  |
|  | Liver | 0 | 9.1 | 10 |  |
|  | Breast | 10 | 18.2 | 10 |  |
|  | Neuroendocrine | 10 | 0 | 0 |  |
|  | Ovary | 10 | 18.2 | 0 |  |
|  | Pancreas | 10 | 9.1 | 10 |  |
|  | Lung | 10 | 0 | 10 |  |
|  | Others | 10 | 0 | 20 |  |
| Chemotherapy | % | 100 | 100 | 100 | 1 |
| Radiotherapy | % | 20 | 12.5 | 0 | 0.594 |

BMI, body mass index. Values are expressed as mean ± standard deviation

The average body mass index (BMI) was 22.1 ± 3.3 kg/m^2^, indicating that the patients were within the normal weight range. However, the weight loss in the last six months was −7.8 ± 6.9 %, with no significant differences between treatment groups (p = 0.891). The most prevalent cancer type was colorectal cancer followed by breast, lung, pancreas and liver cancers with no significant differences between treatments. Treatment adherence was adequate (85.6 %) with no significant difference between treatments (p = 0.337).

### 3.2 Miraculin-Based Food Supplement Efficacy

#### 3.2.1 The effect on electrical and chemical taste perception

Overall, the electrical taste perception did not show significant changes depending on treatment, time, and their interaction with treatment per time (**Table 3**, **Figure 3**). However, patients consuming the standard dose of DMB had the lowest detection levels at the end of the intervention and considerably reduced the taste threshold for an electric‒induced taste stimulus (taste acuity) over time (% change right/left side: ‒52.8 ± 38.5 / ‒ 58.7 ± 69.2 %). None of the cancer patients reached normal thresholds once the intervention was completed (< 7 dB).

**Figure 3.**
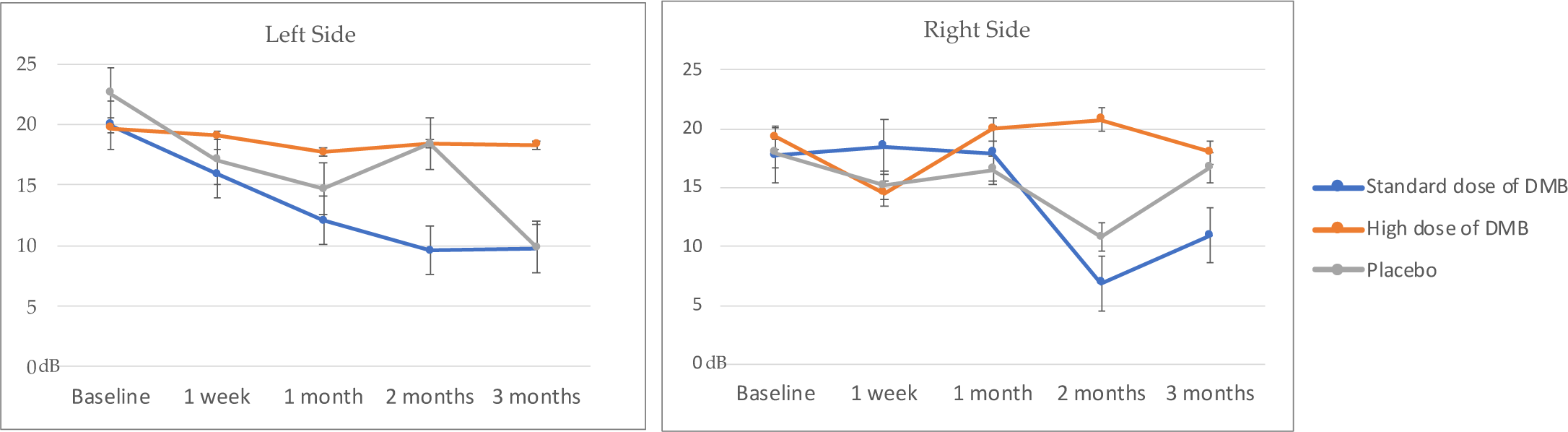
Left and right electrical taste perception (mean ± standard error).

**Table 3.** Electrical taste perception depending on treatment.

|  |  |  |  |  |  | p-value |  |  |
| --- | --- | --- | --- | --- | --- | --- | --- | --- |
|  |  |  | Standard dose of DMB | High dose of DMB | Placebo | Time (t) | Treatment (T) | T × t |
| Right side | (dB) | Baseline | 17.7 ± 13.2 | 19.3 ± 14.0 | 17.9 ± 13.4 | 0.200 | 0.393 | 0.499 |
|  |  | 1 week | 18.5 ± 10.4 | 14.5 ± 15.5 | 15.2 ± 13.5 |  |  |  |
|  |  | 1 month | 17.9 ± 16.3 | 20.0 ± 15.4 | 16.5 ± 17.3 |  |  |  |
|  |  | 2 months | 6.9 ± 10.8 | 20.8 ± 14.1 | 10.8 ± 11.9 |  |  |  |
|  |  | 3 months | 10.9 ± 11.1 | 18.0 ± 18.8 | 16.7 ± 17.1 |  |  |  |
| Left side | (dB) | Baseline | 20.0 ± 12.5 | 19.7 ± 14.0 | 22.6 ± 13.8 | 0.444 | 0.544 | 0.946 |
|  |  | 1 week | 15.9 ± 12.9 | 19.1 ± 16.0 | 17.1 ± 15.8 |  |  |  |
|  |  | 1 month | 12.1 ± 15.3 | 17.7 ± 15.1 | 14.7 ± 15.4 |  |  |  |
|  |  | 2 months | 9.6 ± 13.5 | 18.4 ± 16.2 | 18.4 ± 13.1 |  |  |  |
|  |  | 3 months | 9.8 ± 13.5 | 18.3 ± 18.4 | 9.9 ± 12.5 |  |  |  |

Values are expressed as mean ± standard deviation.

However, at the end of the study, the chemical taste perception reached normal levels (≥ 9) in all patients (**Table 4**). When different tastes were evaluated, salty taste perception changed over time and depending on the treatment assigned (p < 0.001). In this regard, patients consuming DMB significantly improved the perception of salty taste versus placebo (p < 0.05) (**Figure 4**).

**Figure 4.**
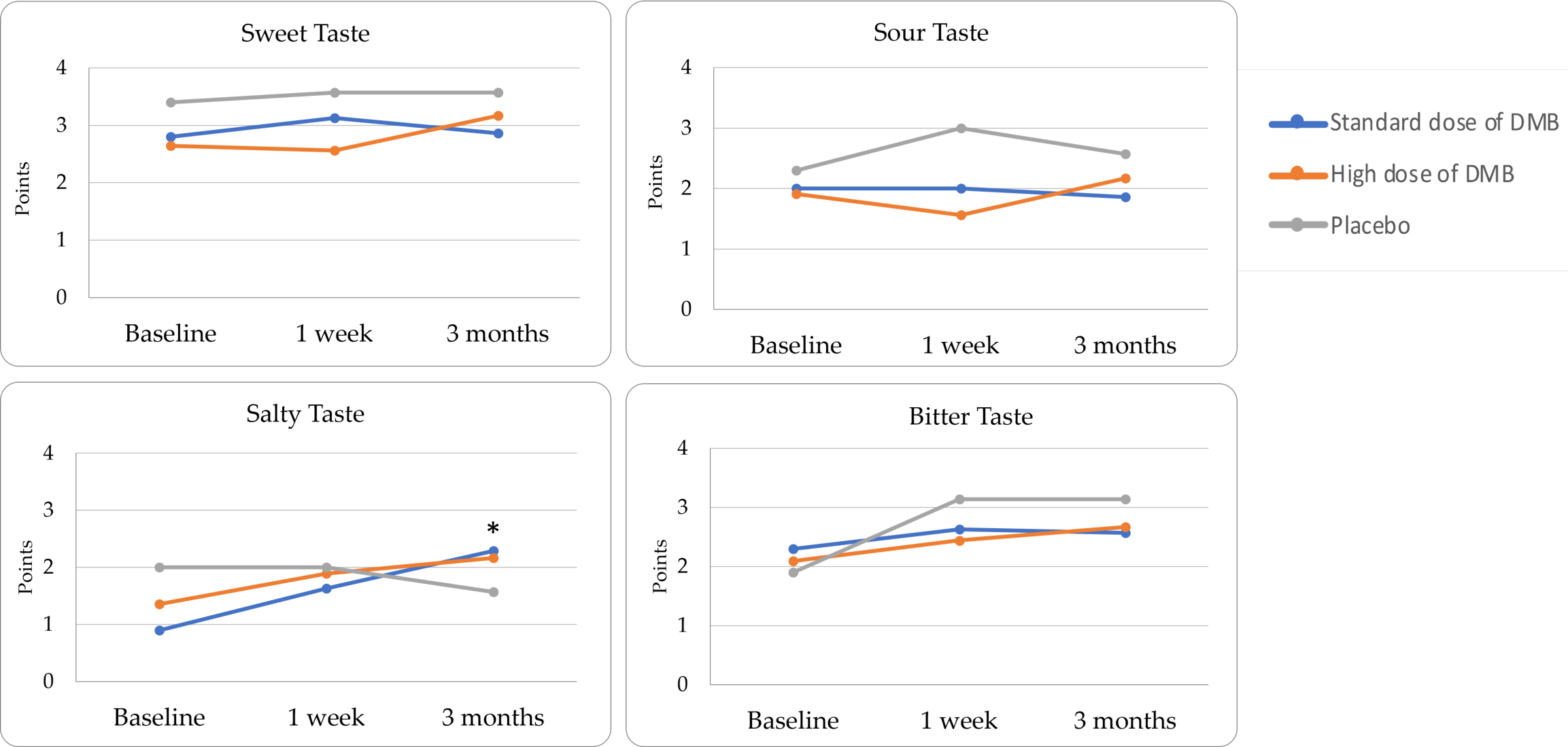
Chemical taste perception (mean ± standard error)

**Table 4.**
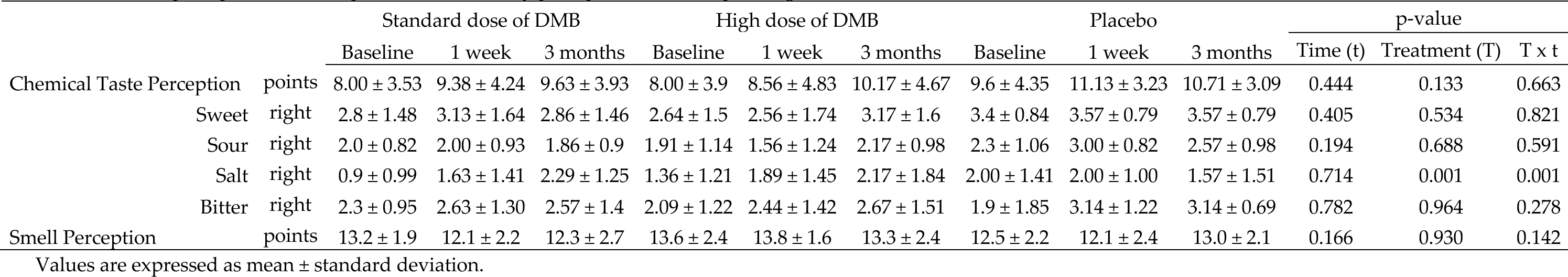
Chemical perception (taste strips test) and olfactory perception (smell) depending on the treatment.

|  |  | Standard dose of DMB |  |  | High dose of DMB |  |  | Placebo |  |  | p-value |  |  |
| --- | --- | --- | --- | --- | --- | --- | --- | --- | --- | --- | --- | --- | --- |
|  |  | Baseline | 1 week | 3 months | Baseline | 1 week | 3 months | Baseline | 1 week | 3 months | Time (t) | Treatment (T) | T × t |
| Chemical Taste Perception | points | 8.00 ± 3.53 | 9.38 ± 4.24 | 9.63 ± 3.93 | 8.00 ± 3.9 | 8.56 ± 4.83 | 10.17 ± 4.67 | 9.6 ± 4.35 | 11.13 ± 3.23 | 10.71 ± 3.09 | 0.444 | 0.133 | 0.663 |
|  | Sweet right | 2.8 ± 1.48 | 3.13 ± 1.64 | 2.86 ± 1.46 | 2.64 ± 1.5 | 2.56 ± 1.74 | 3.17 ± 1.6 | 3.4 ± 0.84 | 3.57 ± 0.79 | 3.57 ± 0.79 | 0.405 | 0.534 | 0.821 |
|  | Sour right | 2.0 ± 0.82 | 2.00 ± 0.93 | 1.86 ± 0.9 | 1.91 ± 1.14 | 1.56 ± 1.24 | 2.17 ± 0.98 | 2.3 ± 1.06 | 3.00 ± 0.82 | 2.57 ± 0.98 | 0.194 | 0.688 | 0.591 |
|  | Salt right | 0.9 ± 0.99 | 1.63 ± 1.41 | 2.29 ± 1.25 | 1.36 ± 1.21 | 1.89 ± 1.45 | 2.17 ± 1.84 | 2.00 ± 1.41 | 2.00 ± 1.00 | 1.57 ± 1.51 | 0.714 | 0.001 | 0.001 |
|  | Bitter right | 2.3 ± 0.95 | 2.63 ± 1.30 | 2.57 ± 1.4 | 2.09 ± 1.22 | 2.44 ± 1.42 | 2.67 ± 1.51 | 1.9 ± 1.85 | 3.14 ± 1.22 | 3.14 ± 0.69 | 0.782 | 0.964 | 0.278 |
| Smell Perception | points | 13.2 ± 1.9 | 12.1 ± 2.2 | 12.3 ± 2.7 | 13.6 ± 2.4 | 13.8 ± 1.6 | 13.3 ± 2.4 | 12.5 ± 2.2 | 12.1 ± 2.4 | 13.0 ± 2.1 | 0.166 | 0.930 | 0.142 |

Values are expressed as mean ± standard deviation.

Particularly, those taking both DMB standard and high doses experienced a notable percentage of change from baseline (108.3 ± 134.4 and 158.3 ± 116.7 %) contrasting with placebo (‒22.2 ± 72.0 %). Although no significant changes were observed depending on time or treatment, bitter taste, frequently affected by chemotherapy treatment, had a lower percentage change in those patients receiving the standard dose of DMB (% change = 14.3 ± 65.6 %) contrasting with the high dose of DMB (25.0 ± 16.7 %) or placebo (33.3 ± 94.3 %). Smell perception did not change throughout the clinical trial (**Table 4**).

#### 3.2.2 The effect on dietary intake

Since the beginning of the study, the diet of the cancer patients was high-protein and high-fat, and this condition persisted during the study. Patients consuming the standard dose of DMB declared not having consumed a smaller amount of food (p = 0.032) considering 22% perceived eating less at the beginning of the study (**Table S1**).

Related to the above, changes in energy intake (p = 0.075) were observed in patients depending on treatment (**Table 5**). Indeed, at the end of the intervention, the group receiving the standard dose of DMB exhibited the highest energy intake compared with the other two groups. Moreover, patients consuming the standard dose of DMB were those who best covered energy expenditure (107 ± 19 %).

**Table 5.** Diet characteristics depending on the assigned treatment.

|  |  | Standard dose of DMB |  |  |  | High dose of DMB |  |  |  | Placebo |  |  |  | p-value |  |  |
| --- | --- | --- | --- | --- | --- | --- | --- | --- | --- | --- | --- | --- | --- | --- | --- | --- |
|  |  | Baseline | 1 month | 2 months | 3 months | Baseline | 1 month | 2 months | 3 months | Baseline | 1 month | 2 months | 3 months | Time (t) | Treatment (T) | T × t |
| Intake | kcal/d | 2512 ± 569 | 2641 ± 384 | 2364 ± 594 | 2679 ± 625 | 2254 ± 663 | 2030 ± 577 | 1809 ± 291 | 1850 ± 778 | 2338 ± 724 | 2294 ± 751 | 2035 ± 301 | 2443 ± 581 | 0.290 | 0.075 | 0.907 |
| Contribution | % | 100 ± 22 | 101 ± 26 | 90 ± 25 | 107 ± 19 | 83 ± 20 | 69 ± 22 | 62 ± 12 | 61 ± 29 | 89 ± 29 | 90 ± 30 | 79 ± 13 | 93 ± 26 | 0.513 | 0.324 | 0.982 |
| Calorie profile |  |  |  |  |  |  |  |  |  |  |  |  |  |  |  |  |
| Proteins | % | 15.8 ± 2.2 | 16 ± 3.2 | 16.7 ± 2.0 | 16.1 ± 1.5 | 16.5 ± 1.72 | 19.4 ± 5.3 | 18.3 ± 2.2 | 18.1 ± 3.4 | 18.6 ± 3.5 | 17.4 ± 2.2 | 18.2 ± 2.0 | 15.6 ± 2.6 | 0.113 | 0.332 | 0.164 |
| Carbohydrates | % | 37.2 ± 4.8 | 34.6 ± 2.9 | 34.2 ± 7.1 | 30.4 ± 4.5 | 37.7 ± 7.88 | 36.0 ± 11.1 | 40.4 ± 9.0 | 39.3 ± 4.8 | 36.0 ± 5.4 | 40.2 ± 3.4 | 36.2 ± 6.1 | 37.5 ± 6.3 | 0.208 | 0.806 | 0.060 |
| Lipids | % | 41.7 ± 4.5 | 44.0 ± 5.3 | 45.3 ± 4.1 | 48.37 ± 5.0 | 42.8 ± 6.81 | 42.0 ± 8. | 37.8 ± 10.2 | 39.5 ± 5.3 | 43.3 ± 6.0 | 39.4 ± 4.4 | 43.1 ± 6.7 | 43.4 ± 3.4 | 0.163 | 0.431 | 0.017 |
| Lipidic profile |  |  |  |  |  |  |  |  |  |  |  |  |  |  |  |  |
| SFA | % | 13.8 ± 2.0 | 12.7 ± 2.4 | 13.4 ± 2.5 | 14.9 ± 3.3 | 12.8 ± 2.7 | 14.3 ± 4.7 | 11.8 ± 3.9 | 12.3 ± 4.0 | 13.5 ± 2.2 | 11.8 ± 3.5 | 13.7 ± 3.7 | 12.0 ± 3.1 | 0.483 | 0.799 | 0.042 |
| MUFA | % | 18.2 ± 4.3 | 20.9 ± 3.5 | 20.4 ± 4.2 | 23.1 ± 4.4 | 18.9 ± 4.3 | 17.5 ± 4.9 | 17.2 ± 6.8 | 16.3 ± 4.3 | 19.4 ± 5.2 | 18.5 ± 3.8 | 19.6 ± 3.8 | 21.0 ± 3.7 | 0.308 | 0.401 | 0.092 |
| PUFA | % | 7.8 ± 9.6 | 41.8 ± 109.1 | 31.1 ± 60.1 | 22.9 ± 46.3 | 29.9 ± 68.9 | 46.8 ± 111.8 | 53.7 ± 120.9 | 29.9 ± 59.1 | 18.8 ± 38.7 | 13.4 ± 24.8 | 14.1 ± 23.7 | 29.0 ± 57.5 | 0.849 | 0.587 | 0.590 |

SFA, Saturated fatty acids; MUFA, Monounsaturated fatty acids; PUFA, Polyunsaturated fatty acids. Values are expressed as mean ± standard deviation.

The energy contribution of lipids (p = 0.017) and carbohydrates (p = 0.060) changed over time and depending on the treatment assigned. Only patients consuming the standard dose of DMB reduced the energy contribution of carbohydrates (% change = –17.6 ± 13.1). Also, these patients had a greater lipid contribution compared to those consuming the high dose of DMB (p = 0.003) or placebo (p = 0.020). In addition, patients taking the standard dose of DMB also had a greater lipid percentage of change from the beginning to the end of the intervention (22.0 ± 15.7 %). Moreover, there was a significant change over time and depending on treatment in the dietary percentage provided by saturated fatty acids (SFA, p = 0.042) and monounsaturated fatty acids (MUFA, p = 0.092). In this regard, patients consuming a standard dose of DMB tended to intake more SFA versus placebo (p = 0.071). Additionally, patients consuming the standard dose of DMB increased all major dietary fatty acids from the beginning to the end of the intervention, including SFA (% change = 11.2 ± 20.2 %), MUFA (40.6 ± 33.2) and polyunsaturated fatty acids (PUFA, 41.1 ± 123.9 %) different from those consuming high dose of DMB (−8.6 ± 32.7; −6.4 ± 15.4; 3.2 ± 45.6 %) or placebo (−12.0 ± 16.5; 4.1 ± 20.6; 1.9 ± 29.9 %).

Taking the latter into account, after three months of intervention, all patients showed a trend to decrease in levels of palmitic and stearic acid from the fatty acid profile of erythrocytes (p = 0.068), particularly in DMB patients (% change standard dose: −13.2 ± 44.0/ −7.9 ± 17.5; high dose: −7.9 ± 29.1/-11.9 ± 19.7; placebo: 3.6 ± 35.9/-1.0 ± 42.3) (**Table 6**, **Figure 5**). In patients consuming standard and high doses of DMB, the increase in linoleic acid percentage change was 15.3 ± 15.0 and 4.7 ± 17.3 % respectively, while it was reduced in placebo (−6.0 ± 20.7 %).

**Figure 5.**
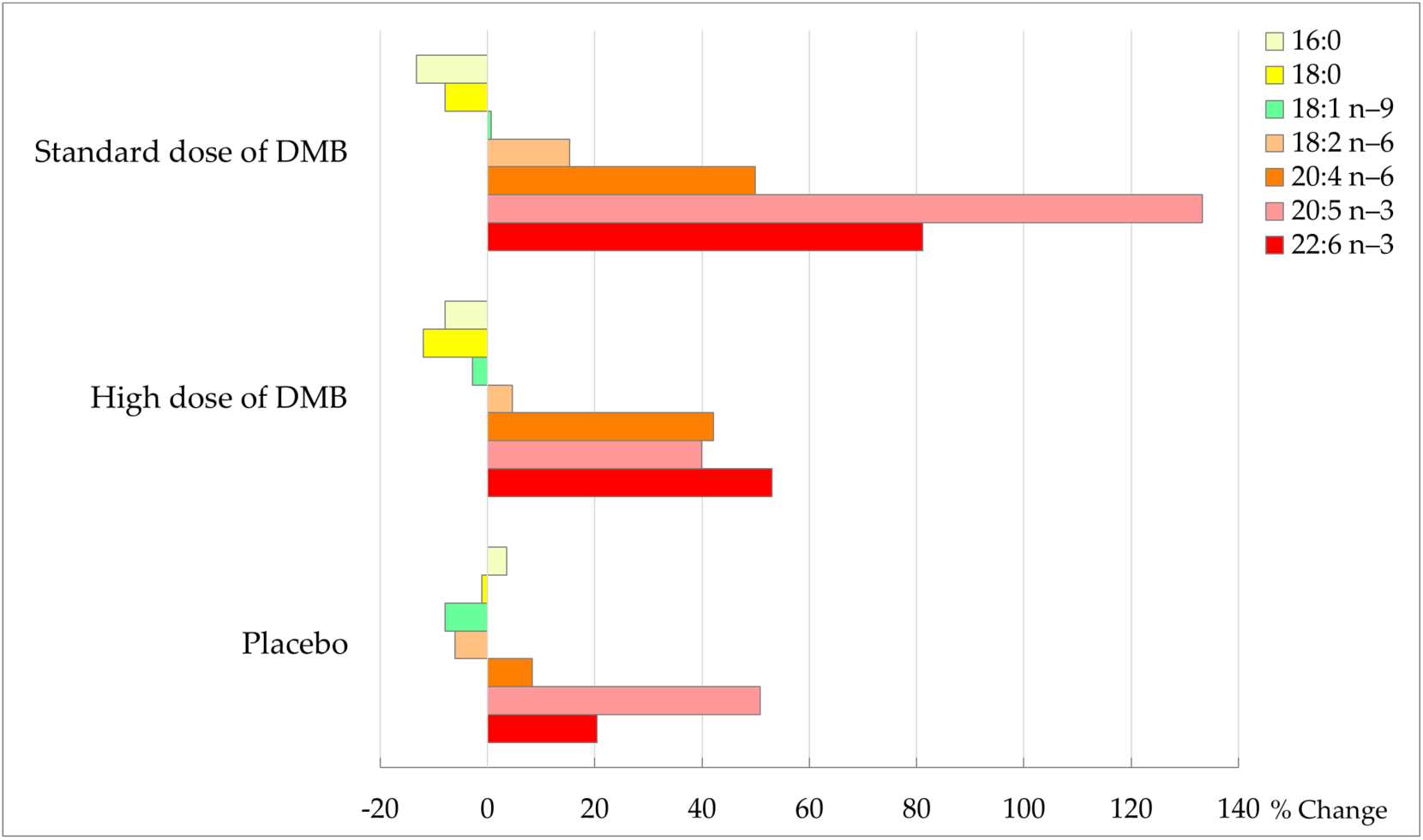
Percentage of change in membrane fatty acids at the end of the intervention (%).

**Table 6.** Fatty acid profile of erythrocytes depending on treatment.

|  |  | Standard dose of DMB |  | High dose of DMB |  | Placebo |  | p-value |  |  |
| --- | --- | --- | --- | --- | --- | --- | --- | --- | --- | --- |
|  |  | Baseline | 3 months | Baseline | 3 months | Baseline | 3 months | Time (t) | Treatment (T) | T × t |
| Palmitic acid (16:0) | % | 26.4 ± 1.8 | 22.4 ± 11.3 | 29.7 ± 8.3 | 26.5 ± 2.6 | 26.3 ± 2.7 | 26.1 ± 7.5 | 0.342 | 0.483 | 0.814 |
| Stearic acid (18:0) | % | 20.4 ± 1.4 | 18.2 ± 2.7 | 21.8 ± 4.6 | 18.6 ± 2.5 | 21.1 ± 3.0 | 19.8 ± 4.3 | 0.068 | 0.676 | 0.817 |
| Oleic acid (18:1 n-9) | % | 19.7 ± 3.1 | 20.0 ± 2.1 | 18.7 ± 3.2 | 17.8 ± 2.8 | 18.2 ± 1.4 | 17.1 ± 1.2 | 0.497 | 0.122 | 0.787 |
| Total PUFA | % | 41.2 ± 1.4 | 47.1 ± 5.2 | 40.7 ± 4.6 | 45.2 ± 5.9 | 41.3 ± 2.8 | 41.9 ± 3.1 | 0.009 | 0.759 | 0.784 |
| Linoleic acid (18:2 n-6) | % | 8.0 ± 0.8 | 9.0 ± 0.8 | 8.2 ± 0.6 | 8.6 ± 1.7 | 8.8 ± 0.9 | 8.4 ± 2.0 | 0.327 | 0.949 | 0.296 |
| Arachidonic acid (20:4 n-6) | % | 10.0 ± 2.8 | 13.1 ± 1.8 | 10.3 ± 2.1 | 14.0 ± 2.8 | 11.1 ± 2.0 | 12.0 ± 2.0 | 0.004 | 0.810 | 0.396 |
| Eicosapentaenoic acid (20:5 n-3) | % | 0.4 ± 0.2 | 0.6 ± 0.4 | 0.5 ± 0.1 | 0.7 ± 0.4 | 0.3 ± 0.2 | 0.5 ± 0.3 | 0.093 | 0.536 | 0.963 |
| Docosahexaenoic acid (22:6 n-3) | % | 3.0 ± 1.3 | 4.4 ± 1.7 | 3.0 ± 1.1 | 4.1 ± 1.0 | 2.9 ± 0.7 | 3.9 ± 1.6 | 0.014 | 0.923 | 0.836 |
| Omega-3 Index | % | 3.4 ± 1.3 | 5.0 ± 2.1 | 3.5 ± 1.2 | 4.8 ± 1.2 | 3.2 ± 0.9 | 4.4 ± 1.4 | 0.010 | 0.936 | 0.947 |

Values are expressed as mean ± standard deviation.

Moreover, there was a change in total PUFA (p = 0.009), total PUFA n‒6 (p = 0.010), arachidonic acid (20:4 n−6, p = 0.004), EPA (20:5 n−3, p = 0.093), DHA (22:6 n−3, p = 0.014) and Omega‒3 index (p = 0.010) over time. In the groups consuming standard and high doses of DMB, AA increased the percentage change by 49.9 ± 57.9 and 42.1 ± 49.8 %, respectively, while in placebo 8.4 ± 31.5 %. The percentage of change of n‒6 PUFA was higher in patients consuming the standard dose of DMB (30.2 ± 26.0 %) and high dose of DMB (23.5 ± 28.5 %) in contrast to placebo (1.5 ± 26.6 %). It was also the standard dose of DMB consumed by cancer patients who observed a greater percentage of change in DHA (81.2 ± 94.7 %) and omega‒3 index (52.7 ± 81.0 %) from the beginning to the end of the intervention.

#### 3.2.3 The effect on anthropometry and body composition

After three months of intervention with the miraculin-based food supplement, oncologic patients tended to change body weight (p = 0.073), BMI (p = 0.073), and waist circumference (p = 0.053) and significantly changed fat-free mass (p = 0.006) and total water (p = 0.029) over time and depending on treatment (**Table 7**). Patients consuming the standard dose of DMB were those who had a higher percentage of change in body weight (‒1.9 ± 4.4 %), BMI (‒1.4 ± 4.6 %) and waist circumference (−2.4 ± 6.7 %) compared to the beginning of the intervention. Compared to placebo, patients consuming the standard dose of DMB increased fat-free mass (p = 0.007) and those with the high dose of DMB had greater total water (p = 0.020). Only patients consuming DMB reduced fat mass, mainly those with a standard dose [(‒2.5 ± 1.3 vs. –1.3 ± 3.2 vs. 0.5 ± 0.8 kg); (% change = ‒11.4 ± 35.0 vs. −6.1 ± 19.5 vs. 2.1 ± 14.8 %)] (**Table 7**).

**Table 7.**
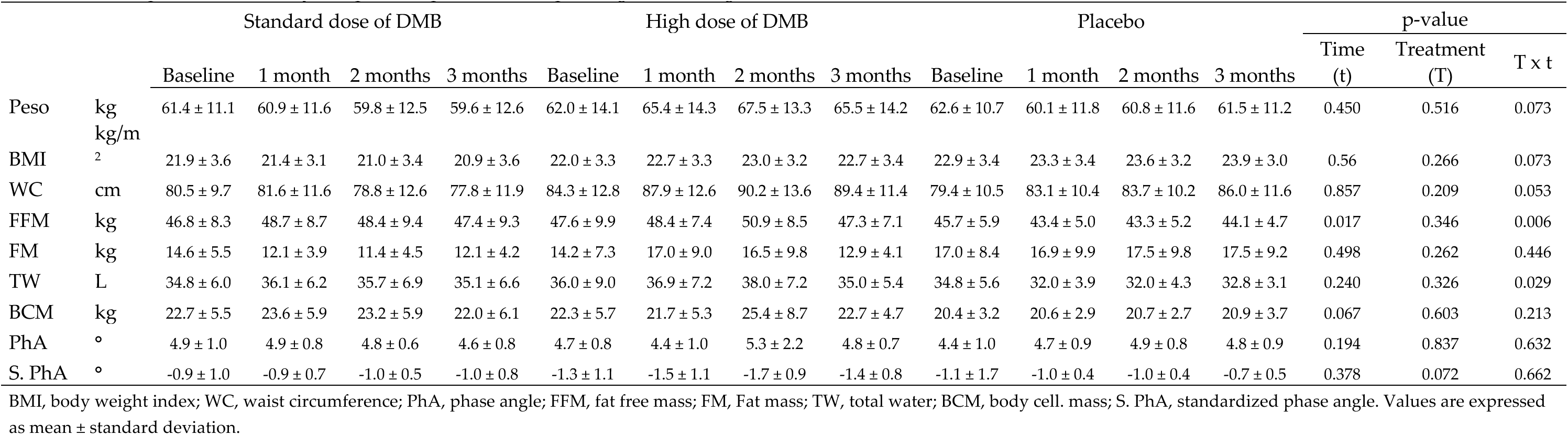
Anthropometric and body composition parameters depending on the assigned treatment.

When the bioimpedance phase angle was evaluated, all patients showed a loss of cellular integrity throughout the study (< 5°). However, when the angle phase was standardized by age and sex, patients treated with DMB tended to present an improvement depending on treatment (p = 0.072). Also, the percentage of change was greater in patients consuming the standard dose of DMB (61.8 ± 19.1 %) than in those with a high dose of DMB (53.7 ± 99.6 %) or placebo, where it worsened (‒20.6 ± 95.6 %) (**Table 7**).

After three months of intervention, all patients regained part of the weight lost during the last 6 months before the start of the study and improved their nutritional status without significant differences between treatments (**Table S2**). Two patients consuming a standard dose of DMB continued with severe malnutrition after the study ended.

#### 3.2.4 The effect on quality of life

Although the global health status perception was not modified by the consumption of the miraculin-based food supplement, changes were observed on social (p = 0.018), fatigue (p = 0.044) and constipation (p = 0.048) scales depending on treatment (**Table 8**). At the end of the intervention patients consuming a high dose of DMB significantly reduced their social scale (p < 0.05) and felt more fatigue (p < 0.05) compared to a standard dose of DMB and placebo. Patients consuming a standard dose of DMB significantly improved the presence of constipation compared to the other two groups (p < 0.05). In this regard, cancer patients consuming the standard dose of DMB showed a higher percentage change in the social functional scale (19.6 ± 40.3 %) and constipation (−66.7 ± 57.7 %) from the beginning to the end of the intervention.

**Table 8.** Quality of life depending on the assigned treatment.

|  |  | Standard dose of DMB |  |  |  | High dose of DMB |  |  |  | Placebo |  |  |  | p-value |  |  |
| --- | --- | --- | --- | --- | --- | --- | --- | --- | --- | --- | --- | --- | --- | --- | --- | --- |
|  |  | Baseline | 1 month | 2 months | 3 months | Baseline | 1 month | 2 months | 3 months | Baseline | 1 month | 2 months | 3 months | Time (t) | Treatment (T) | T x t |
| Global health status | Points | 66.67 ± 12.42 | 66.67 ± 21.65 | 68.75 ± 13.91 | 69.79 ± 16.02 | 46.21 ± 15.97 | 41.67 ± 22.27 | 44.05 ± 19.07 | 40.28 ± 8.19 | 57.41 ± 23.36 | 73.81 ± 23.78 | 73.81 ± 16.27 | 78.57 ± 9.45 | 0.000 | 0.473 | 0.181 |
| **Functional Scales** |  |  |  |  |  |  |  |  |  |  |  |  |  |  |  |  |
| Physical | Points | 94.67 ± 6.89 | 95.56 ± 6.67 | 95.83 ± 7.07 | 94.17 ± 7.51 | 90.3 ± 8.62 | 85.83 ± 12.57 | 89.52 ± 13.25 | 92.22 ± 6.55 | 97.04 ± 4.84 | 96.19 ± 7.56 | 99.05 ± 2.52 | 98.1 ± 5.04 | 0.083 | 0.117 | 0.228 |
| Daily activities | Points | 96.67 ± 7.03 | 98.15 ± 5.56 | 97.92 ± 5.89 | 97.92 ± 5.89 | 90.91 ± 11.46 | 89.58 ± 15.27 | 95.24 ± 8.13 | 91.67 ± 13.94 | 96.3 ± 7.35 | 92.86 ± 8.91 | 95.24 ± 8.13 | 100.00 ± 0.0 | 0.159 | 0.125 | 0.408 |
| Emotional | Points | 89.17 ± 14.19 | 89.81 ± 14.3 | 91.67 ± 11.79 | 89.58 ± 11.57 | 75.76 ± 19.88 | 70.83 ± 17.82 | 70.24 ± 20.33 | 61.11 ± 20.18 | 85.19 ± 15.47 | 91.67 ± 12.73 | 89.29 ± 16.47 | 82.14 ± 26.1 | 0.074 | 0.273 | 0.947 |
| Cognitive | Points | 93.33 ± 11.65 | 94.44 ± 11.79 | 97.92 ± 5.89 | 97.92 ± 5.89 | 80.3 ± 14.56 | 70.83 ± 21.36 | 73.81 ± 16.27 | 69.44 ± 12.55 | 85.19 ± 22.74 | 90.48 ± 13.11 | 90.48 ± 18.9 | 92.86 ± 13.11 | 0.010 | 0.586 | 0.427 |
| Social | Points | 75 ± 22.57 | 88.89 ± 16.67 | 87.5 ± 14.77 | 87.5 ± 17.25 | 60.61 ± 30.98 | 68.75 ± 41.25 | 71.43 ± 34.31 | 58.33 ± 31.18 | 81.48 ± 19.44 | 85.71 ± 15 | 92.86 ± 13.11 | 85.71 ± 20.25 | 0.108 | 0.018 | 0.936 |
| **Symptomatic Scales** |  |  |  |  |  |  |  |  |  |  |  |  |  |  |  |  |
| Fatigue | Points | 24.44 ± 23.89 | 20.99 ± 23.2 | 12.5 ± 12.51 | 13.89 ± 21.21 | 42.42 ± 18.47 | 40.28 ± 18.72 | 44.44 ± 26.45 | 50 ± 26.06 | 38.27 ± 26.12 | 25.4 ± 19.99 | 23.81 ± 23.51 | 20.63 ± 28.28 | 0.092 | 0.044 | 0.307 |
| Nausea and vomiting | Points | 3.33 ± 7.03 | 3.7 ± 11.11 | 2.08 ± 5.89 | 4.17 ± 11.79 | 12.12 ± 18.4 | 10.42 ± 17.68 | 9.52 ± 13.11 | 5.56 ± 13.61 | 7.41 ± 12.11 | 2.38 ± 6.3 | 4.76 ± 8.13 | 0.0 ± 0.0 | 0.457 | 0.232 | 0.517 |
| Pain | Points | 16.67 ± 19.25 | 11.11 ± 18.63 | 8.33 ± 12.6 | 10.42 ± 12.4 | 24.24 ± 31.06 | 35.42 ± 43.13 | 38.1 ± 39.34 | 50 ± 39.44 | 14.81 ± 21.15 | 4.76 ± 12.6 | 7.14 ± 13.11 | 4.76 ± 12.6 | 0.176 | 0.678 | 0.304 |
| Dyspnoea | Points | 3.33 ± 10.54 | 3.7 ± 11.11 | 4.17 ± 11.78 | 0.0 ± 0.0 | 15.15 ± 17.41 | 12.5 ± 24.8 | 4.76 ± 12.6 | 16.67 ± 18.26 | 3.7 ± 11.11 | 4.76 ± 12.6 | 0.0 ± 0.0 | 9.52 ± 25.2 | 0.615 | 0.306 | 0.244 |
| Insomnia | Points | 26.66 ± 30.63 | 11.11 ± 16.67 | 16.67 ± 17.82 | 16.67 ± 25.2 | 24.24 ± 26.21 | 29.16 ± 33.03 | 14.28 ± 26.22 | 22.22 ± 27.21 | 25.92 ± 27.78 | 23.81 ± 25.2 | 28.57 ± 29.99 | 23.81 ± 31.7 | 0.757 | 0.787 | 0.731 |
| Lost of appetite | Points | 6.67 ± 14.05 | 7.41 ± 14.7 | 4.17 ± 11.78 | 4.17 ± 11.78 | 15.15 ± 22.92 | 12.5 ± 17.25 | 14.28 ± 26.22 | 22.22 ± 27.21 | 14.81 ± 24.21 | 38.09 ± 29.99 | 33.33 ± 33.33 | 23.81 ± 31.7 | 0.070 | 0.688 | 0.227 |
| Constipation | Points | 16.67 ± 23.57 | 3.7 ± 11.11 | 4.17 ± 11.78 | 4.17 ± 11.78 | 15.15 ± 27.34 | 0.0 ± 0.0 | 19.05 ± 26.22 | 16.67 ± 18.26 | 18.52 ± 29.39 | 9.52 ± 25.2 | 14.28 ± 17.82 | 9.52 ± 16.26 | 0.608 | 0.048 | 0.716 |
| Diarrhea | Points | 10.00 ± 16.10 | 7.41 ± 14.7 | 8.33 ± 15.43 | 8.33 ± 23.57 | 21.21 ± 26.97 | 29.16 ± 33.03 | 23.81 ± 31.7 | 16.67 ± 18.26 | 14.81 ± 24.21 | 23.81 ± 31.7 | 19.05 ± 26.22 | 23.81 ± 31.7 | 0.174 | 0.723 | 0.879 |
| Financial difficulties | Points | 6.67 ± 14.05 | 3.7 ± 11.11 | 4.17 ± 11.78 | 4.17 ± 11.78 | 15.15 ± 22.92 | 4.17 ± 11.78 | 19.05 ± 26.22 | 11.11 ± 27.21 | 3.7 ± 11.11 | 4.76 ± 12.60 | 4.76 ± 12.60 | 9.52 ± 16.26 | 0.821 | 0.195 | 0.192 |

Values are expressed as mean ± standard deviation.

Over time, a trend of change was also observed on the physical (p = 0.083), emotional (p = 0.074) and loss of appetite (p = 0.070) scales (**Table 8**). Consistent with food consumed perception, those patients consuming the standard dose of DMB showed a higher fall in loss of appetite (% change = ‒100.0 ± 0.0) contrasting with those consuming the high dose (‒33.3 ± 57.7) or placebo (‒33.3 ± 57.7 %) that increased their inappetence. These patients also were the only ones who improved their emotional scale during the intervention (% change = 1.2 ± 9.9 %).

Additionally, to a better quality of life, the perception perceived by cancer patients about product effectiveness tended to improve depending on treatment (p = 0.074) (**Table S3**). Patients consuming the standard dose of DMB showed a better perception of its effectiveness from the start to end of intervention (% chance = 44.2 ± 73.5 vs. 14.6 ± 75.3 or placebo −21.4 ± 44.4 %).

### 3.3 Miraculin-Based Food Supplement Safety

#### 3.3.1 Adverse events

During the study, some adverse events occurred in the patients evaluated (**Table S4**). However, when patients were asked about the possible association with DMB consumption all declared no none of them were associated with these adverse events. Indeed, the *intensit*y of adverse events reported by cancer patients consuming DMB improved once the intervention was completed. In this sense, patients who initially reported a moderate intensity changed from having a moderate intensity to mild or *not described*. Symptoms such as abdominal distention improved only in those patients consuming the standard dose of DMB. When an adverse event occurred, oncologic patients consumed the medication indicated by the physician. Thus, after three months of treatment, patients consuming DMB did not present more adverse events than those consuming placebo.

#### 3.3.2 Biochemical parameters

Glucose metabolism parameters remained within normal ranges in all considered groups (**Table 9**). It is worth mentioning that, in patients consuming the standard dose of DMB, the percentage of change since the beginning of the intervention in insulin concentration was –20.8 ± 39.7 %, while in high dose was ‒1.6 ± 50.2 % and in placebo ‒7.5 ± 23.4 %.

**Table 9.** Parameters of carbohydrate and lipid metabolism and nutritional status depending on the treatment.

|  |  | Standard dose of DMB |  |  |  | High dose of DMB |  |  |  | Placebo |  |  |  | p-value |  |  |
| --- | --- | --- | --- | --- | --- | --- | --- | --- | --- | --- | --- | --- | --- | --- | --- | --- |
|  |  | Baseline | 1 month | 2 months | 3 months | Baseline | 1 month | 2 months | 3 months | Baseline | 1 month | 2 months | 3 months | Time (t) | Treatment (T) | T x t |
| Glucose | mg/dL | 106.1 ± 15.37 | 104.89 ± 12.72 | 100 ± 15.22 | 102.5 ± 13.9 | 108.82 ± 30.68 | 102.13 ± 15.81 | 95 ± 7.98 | 104.83 ± 26.01 | 109.3 ± 22.99 | 104.57 ± 27.96 | 106.86 ± 25.37 | 102.05 ± 23.21 | 0.809 | 0.678 | 0.789 |
| Insulin | μU/mL | 11.3 ± 10.71 | 10.56 ± 8.35 | 6.25 ± 3.33 | 6.21 ± 3.28 | 12.18 ± 7.67 | 9.14 ± 8.11 | 9.29 ± 6.65 | 16 ± 14.46 | 11.3 ± 9.02 | 9 ± 7.44 | 11.29 ± 8.98 | 10.1 ± 6.19 | 0.845 | 0.323 | 0.435 |
| Total Cholesterol | mg/dL | 187.4 ± 33.28 | 174.44 ± 38.76 | 170.63 ± 33.39 | 178.5 ± 27.15 | 174.45 ± 27.19 | 177.13 ± 27.22 | 186.24 ± 23.43 | 188.67 ± 36.46 | 185.2 ± 29.08 | 169.57 ± 26.51 | 183.29 ± 23.62 | 187.1 ± 21.69 | 0.751 | 0.932 | 0.768 |
| HDL Cholesterol | mg/dL | 60.7 ± 23.99 | 53.33 ± 23.08 | 50.88 ± 22.08 | 52.25 ± 25.42 | 53.45 ± 17.95 | 51.63 ± 17.72 | 50.19 ± 19.02 | 56.33 ± 21.71 | 57 ± 24.82 | 55.43 ± 24.58 | 64.14 ± 27.01 | 65.48 ± 28.25 | 0.909 | 0.546 | 0.192 |
| No HDL | mg/dL | 97 ± 0 | 121.11 ± 34.09 | 119.75 ± 31.67 | 131.83 ± 50.7 | 132.5 ± 0 | 125.5 ± 29.77 | 124.14 ± 41.33 | 132.5 ± 0 | 130.75 ± 27.93 | 114.14 ± 15.85 | 118.86 ± 27.27 | 126 ± 21.4 | 0.801 | 0.991 | 0.989 |
| LDL Cholesterol | mg/dL | 105.9 ± 24.12 | 99.67 ± 27.58 | 100.5 ± 26.44 | 106.13 ± 25.93 | 95.91 ± 30.45 | 99.38 ± 30.39 | 107.19 ± 22.49 | 111 ± 27.62 | 100.2 ± 25.19 | 91.29 ± 18.87 | 91.86 ± 19.18 | 97.86 ± 14.68 | 0.244 | 0.721 | 0.694 |
| Triglycerides | mg/dL | 113.8 ± 60.35 | 107.44 ± 49.51 | 96.38 ± 28.87 | 102.42 ± 43.73 | 145.45 ± 67.59 | 134.88 ± 66.2 | 145.38 ± 45.12 | 107.5 ± 26.36 | 140.2 ± 44.86 | 114.29 ± 30.51 | 135.57 ± 60.35 | 120.33 ± 50.46 | 0.203 | 0.58 | 0.506 |
| Total proteins | g/dL | 6.77 ± 0.45 | 6.68 ± 0.33 | 6.78 ± 0.31 | 6.53 ± 0.38 | 6.96 ± 0.7 | 6.79 ± 0.76 | 6.86 ± 0.57 | 6.95 ± 0.62 | 6.2 ± 1.47 | 6.66 ± 0.5 | 6.71 ± 0.34 | 6.79 ± 0.43 | 0.268 | 0.502 | 0.415 |
| Albumin | g/dL | 4.35 ± 0.17 | 4.22 ± 0.2 | 4.31 ± 0.2 | 4.06 ± 0.5 | 4.24 ± 0.31 | 4.1 ± 0.23 | 4.2 ± 0.21 | 4.32 ± 0.23 | 4.33 ± 0.36 | 4.17 ± 0.28 | 4.21 ± 0.23 | 4.27 ± 0.21 | 0.595 | 0.062 | 0.114 |
| Prealbumin | mg/dL | 23.8 ± 7.11 | 22.22 ± 5.47 | 20.47 ± 5.29 | 20.29 ± 5.76 | 22.07 ± 7.96 | 21.49 ± 8.34 | 23.73 ± 8.94 | 24.94 ± 11.42 | 19.94 ± 5.68 | 17.61 ± 3.76 | 19.74 ± 3.34 | 20.58 ± 4.55 | 0.337 | 0.297 | 0.152 |
| RBP | mg/dL | 4.36 ± 1.68 | 3.93 ± 1.47 | 3.79 ± 1.32 | 3.76 ± 1.64 | 4.22 ± 1.34 | 3.99 ± 1.54 | 4.29 ± 1.42 | 4.69 ± 2 | 3.88 ± 1.28 | 3.07 ± 0.6 | 3.33 ± 0.78 | 4.16 ± 1.32 | 0.47 | 0.218 | 0.027 |

RBP, Retinol binding protein. Values are expressed as mean ± standard deviation.

Even though the diet of patients consuming the standard dose of DMB was fat–high (**Table 5**), the blood lipid profile was not altered and parameters related to lipid metabolism remained within normal ranges for the age and sex of the population (**Table 9**).

Proteins usually related to nutritional status such as retinol-binding protein (RBP) showed changes over time and depending on treatment (p = 0.027). Patients consuming the high dose of DMB had higher RBP values than placebo (p < 0.05); however, the mean of this increase remained within normal ranges.

Vitamin and mineral biomarkers, except for magnesium, were not affected by habitual consumption of the miraculin-based food supplement and remained stable throughout the clinical trial and within the normal ranges of the population throughout the clinical trial (**Table S5**). Magnesium showed a change throughout the study depending on the time and treatment assigned (p = 0.028). Only those patients consuming DMB improved magnesium concentration at the end of the study (% change standard dose 4.2 ± 5.7; high dose: 11.7 ± 13.6; placebo −3.0 ± 12.7).

At the end of the study kidney function biomarkers such as creatinine (p = 0.054), glomerular filtration rate (p = 0.051) and uric acid (p = 0.066) tended to change over time and depending on treatment (**Table 10**). Nevertheless, all patients had values within normal ranges.

**Table 10.** Security parameters depending on the assigned treatment.

|  |  | Standard dose of DMB |  |  |  | High dose of DMB |  |  |  | Placebo |  |  |  | p-value |  |  |
| --- | --- | --- | --- | --- | --- | --- | --- | --- | --- | --- | --- | --- | --- | --- | --- | --- |
|  |  | Baseline | 1 month | 2 months | 3 months | Baseline | 1 month | 2 months | 3 months | Baseline | 1 month | 2 months | 3 months | Time (t) | Treatment (T) | T × t |
| Creatinine | mg/dL | 0.76 ± 0.27 | 0.79 ± 0.27 | 0.85 ± 0.26 | 0.77 ± 0.28 | 0.71 ± 0.2 | 0.7 ± 0.18 | 0.68 ± 0.15 | 0.71 ± 0.25 | 1 ± 1.03 | 0.65 ± 0.11 | 0.68 ± 0.12 | 0.75 ± 0.18 | 0.329 | 0.033 | 0.054 |
| GFR | mL/min/1.73 m<sup>2</sup> | 82 ± 13.22 | 79.44 ± 13.74 | 78.38 ± 15.83 | 81.17 ± 14.03 | 87.55 ± 8.68 | 89.63 ± 3.89 | 90.14 ± 2.27 | 87.17 ± 9.39 | 81.1 ± 23.74 | 88.14 ± 7.56 | 86.57 ± 8.42 | 81.71 ± 14.19 | 0.832 | 0.202 | 0.051 |
| Uric acid | mg/dL | 4.25 ± 1.27 | 4.56 ± 1.77 | 4.86 ± 2.08 | 4.12 ± 1.52 | 4.38 ± 1.2 | 4.59 ± 0.82 | 4.64 ± 1 | 4.55 ± 1.22 | 5.1 ± 2 | 4.34 ± 1.13 | 4.77 ± 0.98 | 5.01 ± 1.28 | 0.16 | 0.057 | 0.066 |
| AST | UI/L | 31.64 ± 12.61 | 29.5 ± 18.73 | 27.1 ± 18.42 | 26.39 ± 12.27 | 29.37 ± 17.57 | 30.96 ± 22.07 | 35.86 ± 30.03 | 31.67 ± 21.56 | 26.75 ± 15.23 | 26 ± 10.77 | 28.29 ± 20.31 | 28.05 ± 14.15 | 0.778 | 0.985 | 0.903 |
| ALT | UI/L | 34.2 ± 18.02 | 24.33 ± 12.86 | 27.88 ± 15.01 | 23.88 ± 12.8 | 26.64 ± 13.31 | 25.13 ± 18.91 | 31.19 ± 24.55 | 29.5 ± 13.77 | 32.11 ± 11.6 | 32.71 ± 14.51 | 36.43 ± 16.52 | 37.71 ± 17.98 | 0.221 | 0.051 | 0.057 |
| LDH | UI/L | 225.6 ± 25.68 | 230.75 ± 43.44 | 238.75 ± 27.86 | 229.25 ± 23.75 | 291.8 ± 154.17 | 236.08 ± 77.87 | 272.07 ± 109.24 | 240.08 ± 117.92 | 520.74 ± 841.91 | 258 ± 65.68 | 262 ± 53.59 | 248.11 ± 60.64 | 0.267 | 0.865 | 0.996 |
| AP | UI/L | 82.57 ± 27.28 | 79.17 ± 20.15 | 69.8 ± 9.65 | 72.07 ± 10.39 | 149.6 ± 138.72 | 145.56 ± 172.63 | 167.79 ± 199.69 | 89 ± 31.03 | 146.78 ± 75.96 | 128.14 ± 43.13 | 131.14 ± 46.46 | 126.48 ± 40.81 | 0.264 | 0.934 | 0.982 |
| GGT | UI/L | 39.14 ± 20.31 | 35.33 ± 19.19 | 31 ± 17.07 | 34.07 ± 18.74 | 168.3 ± 389.02 | 235 ± 523.74 | 250 ± 544.6 | 43.17 ± 29.39 | 142.63 ± 157.81 | 92.33 ± 92.83 | 117.67 ± 129.16 | 116.06 ± 122.14 | 0.537 | 0.727 | 0.748 |
| Bilirubin | mg/dL | 0.74 ± 0.45 | 0.66 ± 0.37 | 0.6 ± 0.24 | 0.65 ± 0.36 | 0.53 ± 0.13 | 0.52 ± 0.3 | 0.47 ± 0.24 | 0.46 ± 0.15 | 0.56 ± 0.31 | 0.43 ± 0.2 | 0.52 ± 0.19 | 0.5 ± 0.22 | 0.239 | 0.730 | 0.867 |

GFR, glomerular filtration rate; AST, aspartate aminotransferase; ALT, alanine transaminase; LDH, lactate dehydrogenase; AP, alkaline phosphatase; GGT, gamma-glutamyl transferase. Values are expressed as mean ± standard deviation.

Finally, safety biomarkers of liver function did not show significant changes after completing the clinical trial (**Table 10**), except for ALT levels (p = 0.057). Only patients consuming the standard dose of DMB reduced ALT levels from the beginning to the end of the intervention (% change = ‒7.5 ± 23.4 %, high dose: 16.7 ± 32.9 % and placebo: 5.6 ± 23.2 %) within the normal range while this was not so in patients consuming the placebo who had final ALT blood concentrations higher than normal (< 35 UI/L). From the beginning of the intervention to the end, lactate dehydrogenase (LDH), an enzyme used to detect tissue or liver damage, had higher levels than those recommended (100–190 UI/L) in all patients.

It is worth mentioning that, although there were no differences depending on time or treatment, at the end of the intervention the gamma-glutamyl transferase (GGT), a biomarker of possible damage to the bile ducts, was normal just in those patients consuming the standard dose of DMB while the rest were above normal ranges (< 38 IU/L).

## 4. Discussion

The main findings of the present study were that the habitual intake of a standard dose of DMB improved the electrochemical perception of taste in cancer patients allowing a greater food intake and a better quantity and quality of dietary lipid intake, which in turn was reflected in an ameliorated fatty acids status. Additionally, improvements in body composition, nutritional status and quality of life were observed. Furthermore, the main safety parameters remained stable and within normal ranges throughout the entire study. These results suggest that habitual consumption of a standard dose of 150 mg of miraculin–food supplement (DMB) is effective and safe for malnourished cancer patients in active treatment who present an objective TDs.

Two clinical trials have been carried out on patients receiving chemotherapy using the miracle berry. In the first study, a crossover clinical trial was carried out on 23 chemotherapy patients whose taste alterations were measured by the Wickham questionnaire [39]. In two weeks, patients consumed either the miracle fruit or supportive measures alone. At the end of the study, 30% of patients showed an improvement in taste. The second study included eight participants who received three or more cycles of chemotherapy and expressed positive taste changes to the nurse [40]. These patients were assigned to the experimental (n = 4) or control group (n = 4) in a nonrandomized manner. Patients consumed six fruits per day of miracle fruit or dried cranberries as a placebo for two weeks. At the end of the study, all patients reported positive taste changes with miracle fruit consumption through qualitative data.

In the present study, a reduction in the electrical threshold (taste acuity) was observed in all patients evaluated, including those consuming the miraculin-based food supplement. This finding is relevant because a gradual deterioration in taste perception is expected to occur because of antineoplastic treatment [3,4,49,50] and this deterioration has remained stable throughout the study. Although the overall change in electrical taste perception change was not conclusive, the chemical perception of salty taste significantly improved in cancer patients habitually consuming the standard dose of DMB. Analysis of subjective taste changes reported that salt and umami tastes are more sensitive to chemotherapy than other taste descriptors [51]. Salty taste distortion is the most frequently reported taste alteration during neo/adjuvant chemotherapy [52]. Umami taste was not evaluated as a descriptor in the present clinical trial because foods providing umami flavor are not commonly used in the Spanish population. Since one taste perception is associated with changes in other tastes during chemotherapy [53] an improvement in an affected descriptor can contribute to a better perception of global food taste.

Up to 87% of cancer patients with TDs experience a loss of appetite [54] which is widely known to be associated with poor prognosis [55]. However, patients who consumed the standard dose of DMB did not exhibit a loss of appetite at the end of the study. Therefore, habitual consumption of a standard dose of DMB may protect against loss of appetite in cancer patients; in fact, these patients had greater food intake and better met their energy needs. This finding is of relevance since cancer patients have shown a lower intake of total energy, protein and fat during chemotherapy related to TDs [56].

In addition to better covering the total energy expenditure, habitual consumption of a standard dose of DMB was associated with increased quantity and quality of fat intake in cancer patients. Various studies have shown that high-fat diets, especially those rich in trans and saturated fat, promote tumorigenesis by modulating the gut microbiota [57–59], systemic low-grade inflammation [60], and changes in the adipocytokine profile [61,62]. On the other hand, although epidemiological data do not support the theory that a decrease in total fat intake is effective in preventing cancer [63–66] or decreasing cancer-specific mortality [67], dietary lipid composition can have an impact on cancer pathogenesis [68]. Thus, cancer patients who consumed a standard dose of DMB exhibited notably improved MUFA and PUFA intake. MUFA intake has been inversely associated with decreased cancer risk [63,69]. Indeed, a higher intake of MUFA from plant sources was associated with lower mortality rates associated with all causes [70]. Olive oil is the largest contributor to MUFA since it provides up to 78 % of oleic acid, the most abundant MUFA in the Spanish diet [71]. Thus, olive oil was the most commonly used culinary fat by cancer patients in the present study. A meta-analysis of case-control studies showed that olive oil consumption was associated with lower odds of developing any type of cancer [72], which highlights the importance of its consumption.

On the other hand, a majority of studies examining the relationship between PUFAs and cancer risk have focused on n–6 and n–3, two of their most biologically active representatives. However, a meta-analysis of observational studies revealed a mild inverse association between diets high in total PUFA and specific-cancer risk [73], while others have not found an association with increased risk [66,74]. Therefore, an adequate quantity and quality of dietary fats, promoted by the habitual consumption of a standard dose of DMB could improve the prognosis of these patients.

As shown in the present study, erythrocyte percentages of oleic acid and selected PUFA, including linoleic acid, AA, and DHA, increased following habitual intake of a standard dose of DMB. Additionally, cancer patients who consumed DMB had the highest omega-3 index, an indicator of omega-3 status and coronary heart disease risk [75]. A higher omega-3 index has also been found to be inversely associated with lower cancer-specific risk in a meta-analysis of case-control studies [76]. PUFA play important roles as precursors of lipid mediators that regulate metabolic pathways and inflammatory responses, oxidative stress, and modifications of membrane composition that could impact cell signaling pathways and cancer progression [77]. In addition, cancer cells with more membranes are less susceptible to oxidative stress induced by chemotherapeutic agents [78].

On the other hand, cancer patients undergoing chemotherapy often suffer from nutritional alterations, particularly in terms of essential fatty acid and long-chain PUFA status [79]. Additionally, nutritional status is associated with poor prognosis, lower treatment completion and greater healthcare consumption [80]. Accordingly, it has been reported that supplementation with EPA and DHA in cancer patients has a positive impact during treatment, which is associated with cellular membrane modulation [81]. Moreover, the discovery of pro-resolution mediators of inflammation derived from arachidonic acid, called lipoxins, and from EPA and DHA, called resolvins, protectins and maresins [82–84], supports the idea that a PUFA-enriched membrane could be favorable for the management of this disease [85,86]. In this scenario, it is possible to assume that consuming more and better quality food would involve the intake of more essential fatty acids and lead to an improvement in the levels of PUFA with a concomitant improvement in nutritional status [87]. Changes in the fatty acid profile of the erythrocyte membrane would be indicative of improved nutritional status in cancer patients. This improvement can be attributed to supplementation with the miraculin food supplement given that it was extended for 12 weeks, sufficient time for the complete renovation of the total pool of erythrocytes [88].

In a randomized clinical trial carried out on malnourished cancer patients a high–fat diet provided for eight weeks improved weight control, fat–free mass and body mass from the first to the third chemotherapy cycle [89]. In this regard, in the present clinical trial, habitual consumption of a standard dose of DMB maintained body weight and increased fat–free mass, as measured by BIA, a reliable tool in nutritional intervention studies [90]. This is probably because a high–fat diet, favored by the consumption of DMB, would compensate at least in part for the rise in resting energy expenditure observed in cancer patients [66], which is also a major determinant of the development of malnutrition [91]. Calorie intake is also a significant factor in preventing fat–free mass weight loss in cancer patients [92], and those consuming a standard dose of DMB adequately meet their energy requirements.

Malnutrition predicts the risk of physical impairment, chemotherapy toxicity and mortality in cancer patients [93,94]. In this sense, all cancer patients improved their nutritional status once the intervention was completed. Loss of body weight (skeletal muscle and body fat) is associated with a reduction in quality of life [95]. The latter is also affected by the disease itself and the antineoplastic treatment used [96]. Therefore, it is not surprising that poor quality of life in cancer patients is associated with poor nutritional status [97] and conversely, that malnutrition reduces their quality of life [98]. Additionally, quality of life can significantly impact long-term cancer survivorship [99]. In this regard, in the present clinical trial, it was found that habitual consumption of a standard dose of DMB improved quality of life, in particular constipation, as measured by symptom scales, Diverse catabolic factors are activated by the presence of constipation, fatigue, nausea, vomiting and other relevant symptoms usually present in cancer patients [100]. Fatigue or loss of appetite are among the most common symptoms exhibited by cancer patients that affect their quality of life [101]. In the present study, only patients who consumed a standard dose of DMB improved their loss of appetite and improved their scores on the emotional scale from the beginning to the end of the intervention. They also showed improvements in fatigue. Since TDs caused by cancer therapies negatively affect patient quality of life [14,28,54], the improvement observed in the perception of salty taste in patients consuming a standard dose of DMB could have contributed to the improvement of these quality of life scales.

*Synsepalum dulcificum* fruits have been consumed since the 18th century by natives of Western and Central Africa [102] without describing adverse events beyond wanted taste changes. In 2021, DMB obtained from dried fruits of *S. dulcificum* was approved as a *novel food* in the European Union after a positive scientific opinion by the European Food Safety Authority (EFSA). The panel concluded that an intake of 10 mg/kg body weight (bw) per day is safe for human consumption [41]. The maximum dose used in the present clinical trial was 0.9 g/day, slightly above this recommendation. However, the EFSA also indicated that a 90–day oral dose of 2000 mg/kg bw per day was not associated with adverse effects. In this vein, different studies assessed the taste-modifying properties of different products from *S. dulcificum* and although this has not been its main objective, the authors of these studies did not report adverse events during its consumption [40,103–107]. The potential allergenicity and toxicity of miraculin have also been evaluated and it has not been associated with any safety concerns [108].

In this regard, cancer patients who habitually consumed DMB did not experience any adverse events related to their consumption. A negative effect, but not an adverse event, was the dropout of six patients due to the taste distortion caused by habitually non-sweet acidic foods such as tomatoes and salads. The majority of dropouts (67 %) occurred at a high dose of DMB, indicating that patients are more likely to accept a standard dose of DMB. Indeed, the effectiveness perceived by patients of the food supplement containing miraculin increased notably in those patients consuming a standard dose of DMB over time. Several studies have shown that the degree of the taste-modifying effect of miracle berries differs according to fruit type, source or preparation [109] since it determines the miraculin content. The smaller quantity the lower the sweetness intensity and *vice versa* [107]. A high dose of DMB, with a higher miraculin content, probably provided high sweetness intensity and persistence, significantly modifying the cancer patient’s taste of sour foods. This is because miraculin stimulates a sweet taste 400,000 times greater than sucrose [110] and its effect can linger up to two hours until miraculin dissociates from the taste receptors by the action of salivary amylase [111].

While the energetic contribution of dietary lipids increased significantly in those consuming the standard dose of DMB, its continued consumption for 3 months did not alter the blood lipid profile. Triterpenoids isolated from the miracle fruit can act as cholesterol-lowering agents [112] and as effective antihyperglycemic agents [113] by increasing insulin synthesis, inhibiting carbohydrate metabolizing enzymes [114] and improving insulin sensitivity [115]. In this regard, it is worth mentioning that the plasma lipid profile, as well as glucose metabolism parameters, remained stable and within normal ranges throughout the intervention.

Habitual consumption of a standard dose of DMB may have a hepatoprotective effect since the placebo patients had liver markers such as ALT and GGT above normal ranges. The hepatoprotective effect of miracle berries has already been described in previous experimental studies [113]. Kidney protection was also been observed when miracle fruit extracts were used. Indeed, it has been proposed as a novel plasma uric-lowering agent [116]. In this sense, it was observed that patients consuming a standard dose of DMB tended to reduce, within normal ranges, the concentration of uric acid.

The major strength of the present clinical trial was the use of objective analysis in the evaluation of the effect of habitual consumption of a food supplement containing miraculin on electrochemical taste perception in cancer patients undergoing active treatment. Due to the exploratory nature of the present study, one of the limitations was the reduced number of patients evaluated. Additionally, the complexity of managing cancer patients (polypharmacy, complications, intercurrent diseases, etc.) may have conditioned the high treatment dropout rate. However, based on the results obtained at the present study, the calculation of the ideal sample size will allow us to confirm and expand the results in future clinical trials as the DMB optimal dose has now been established.

## 5. Conclusions

Habitual consumption of a standard dose of DMB, equivalent to 150 mg of the dried berry, before each main meal, improves electrochemical food perception allowing greater food intake and a better quantity and quality of the lipid profile reflected in the diet and membrane fatty acids. Additionally, a standard dose of DMB increases fat–free mass and reduces fat mass but also promotes improvements in quality–of–life such as constipation. The nutritional status of cancer patients who consumed a standard dose of DMB also improved. Additionally, the habitual consumption of DMB appears to be safe with no changes in major biochemical parameters associated with health status.

## Supplementary Materials

The following supporting information can be downloaded at: www.mdpi.com/xxx/s1, Table S1: Perception of food consumption depending on treatment; Table S2: Nutritional status depending on treatment; Table S3: Perceived effectiveness of the product depending on treatment; Table S4: Adverse events depending on the assigned treatment group; Table S5: Vitamins and Minerals depending on treatment.

## Author Contributions

Conceptualization, B.L.-P., A.G., and S.P.-M.; methodology, B.L.-P., T.H., and J.F.-B.; software, J.D.-P.; validation, F.J.R.-O, and M.B.-H.; formal data analysis, J.D.-P.; investigation, B.L.-P and L.A.-C.; resources, S.P.-M.; data curation, L.A.-C; writing—original draft preparation, B.L.-P., A.I.A.-M, A.G.; writing—review and editing, B.L.-P and A.G.; visualization, A.I.A.-M.; supervision, S.P.-M and A.G.; project administration, B.L.-P; funding acquisition, S.P.-M. All authors have read and agreed to the published version of the manuscript.

## Funding

This study is funded by Medicinal Gardens S.L. through the Center for Industrial Technological Development (CDTI), “Cervera” Transfer R&D Projects. Ref. IDI-20210622. (Science and Education Ministry, Spain).

## Institutional Review Board Statement

The study was conducted following the Declaration of Helsinki and approved by the Ethics Committee of Hospital Universitario La Paz (protocol code 6164 and Jun 23rd 2022 date of approval).

## Informed Consent Statement

Informed consent was obtained from all subjects involved in the study.

## Data Availability Statement

The data used to support the findings of this study are available from the corresponding author upon request.

## Data Availability

Informed consent was obtained from all subjects involved in the study.

## Acknowledgments

The authors would like to thank Medicinal Gardens S.L. (Baïa Food Co.) for providing the orodispersible DMB^®^ tablets (TasteCare^®^) and for its support and technical advice.

## Conflicts of Interest

The authors declare no conflicts of interest.

## Notes

### Competing Interest Statement

The authors have declared no competing interest.

### Clinical Trial

NCT05486260

## References

1. Gamper, E.M.; Giesinger, J.M.; Oberguggenberger, A.; Kemmler, G.; Wintner, L.M.; Gattringer, K.; Sperner-Unterweger, B.; Holzner, B.; Zabernigg, A. Taste Alterations in Breast and Gynaecological Cancer Patients Receiving Chemotherapy: Prevalence, Course of Severity, and Quality of Life Correlates. Acta Oncol. 2012, 51, 490–496, doi:10.3109/0284186X.2011.633554.

2. Zabernigg, A.; Gamper, E.-M.; Giesinger, J.M.; Rumpold, G.; Kemmler, G.; Gattringer, K.; Sperner-Unterweger, B.; Holzner, B. Taste Alterations in Cancer Patients Receiving Chemotherapy: A Neglected Side Effect? Oncologist 2010, 15, 913–920, doi:10.1634/THEONCOLOGIST.2009-0333.

3. Bernhardson, B.M.; Tishelman, C.; Rutqvist, L.E. Self-Reported Taste and Smell Changes during Cancer Chemotherapy. Support. Care Cancer 2008, 16, 275–283, doi:10.1007/S00520-007-0319-7.

4. Bernhardson, B.M.; Tishelman, C.; Rutqvist, L.E. Chemosensory Changes Experienced by Patients Undergoing Cancer Chemotherapy: A Qualitative Interview Study. J. Pain Symptom Manage. 2007, 34, 403–412, doi:10.1016/J.JPAINSYMMAN.2006.12.010.

5. Heckmann, J.G.; Heckmann, S.M.; Lang, C.J.G.; Hummel, T. Neurological Aspects of Taste Disorders. Arch. Neurol. 2003, 60, 667–671, doi:10.1001/ARCHNEUR.60.5.667.

6. Epstein, J.B.; Barasch, A. Taste Disorders in Cancer Patients: Pathogenesis, and Approach to Assessment and Management. Oral Oncol. 2010, 46, 77–81, doi:10.1016/J.ORALONCOLOGY.2009.11.008.

7. Abasaeed, R.; Coldwell, S.E.; Lloid, M.E.; Soliman, S.H.; Macris, P.C.; Schubert, M.M. Chemosensory Changes and Quality of Life in Patients Undergoing Hematopoietic Stem Cell Transplantation. Support. Care Cancer 2018, 26, 3553–3561, doi:10.1007/S00520-018-4200-7/METRICS.

8. Shah, M.H.; Kloos, R.T.; Ringel, M.D.; Knopp, M. V.; Hall, N.C.; King, M.; Stevens, R.; Liang, J.; Wakely, P.E.; Vasko, V. V.;, et al. Phase II Trial of Sorafenib in Metastatic Thyroid Cancer. J. Clin. Oncol. 2009, 27, 1675–1684, doi:10.1200/JCO.2008.18.2717/ASSET/IMAGES/ZLJ9990984060006.JPEG.

9. Adjei, A.A.; Molina, J.R.; Mandrekar, S.J.; Marks, R.; Reid, J.R.; Croghan, G.; Hanson, L.J.; Jett, A.R.; Xia, C.; Lathia, C.;, et al. Phase I Trial of Sorafenib in Combination with Gefitinib in Patients with Refractory or Recurrent Non–Small Cell Lung Cancer. Clin. Cancer Res. 2007, 13, 2684–2691, doi:10.1158/1078-0432.CCR-06-2889.

10. Buttiron Webber, T.; Briata, I.M.; DeCensi, A.; Cevasco, I.; Paleari, L. Taste and Smell Disorders in Cancer Treatment: Results from an Integrative Rapid Systematic Review. Int. J. Mol. Sci. 2023, 24, doi:10.3390/IJMS24032538.

11. de Vries, Y.C.; Winkels, R.M.; van den Berg, M.M.G.A.; de Graaf, C.; Kelfkens, C.S.; de Kruif, J.T.C.M.; Göker, E.; Grosfeld, S.; Sommeijer, D.W.; van Laarhoven, H.W.M.;, et al. Altered Food Preferences and Chemosensory Perception during Chemotherapy in Breast Cancer Patients: A Longitudinal Comparison with Healthy Controls. Food Qual. Prefer. 2018, 63, 135– 143, doi:10.1016/J.FOODQUAL.2017.09.003.

12. Postma, E.M.; de Vries, C.; Boesveldt, S. Tasty Food for Cancer Patients: The Impact of Smell and Taste Alterations on Eating Behaviour. Ned Tijdschr Geneeskd 2017, 160, D748.

13. Von Grundherr, J.; Koch, B.; Grimm, D.; Salchow, J.; Valentini, L.; Hummel, T.; Bokemeyer, C.; Stein, A.; Mann, J. Impact of Taste and Smell Training on Taste Disorders during Chemotherapy - TASTE Trial. Cancer Manag. Res. 2019, 11, 4493–4504, doi:10.2147/CMAR.S188903.

14. Hovan, A.J.; Williams, P.M.; Stevenson-Moore, P.; Wahlin, Y.B.; Ohrn, K.E.O.; Elting, L.S.; Spijkervet, F.K.L.; Brennan, M.T. A Systematic Review of Dysgeusia Induced by Cancer Therapies. Support. Care Cancer 2010, 18, 1081–1087, doi:10.1007/S00520-010-0902-1.

15. Ghias, K.; Jiang, Y.; Gupta, A. The Impact of Treatment-Induced Dysgeusia on the Nutritional Status of Cancer Patients. 2023, doi:10.1016/j.nutos.2023.06.004.

16. Brand, J.G. Within Reach of an End to Unnecessary Bitterness? Lancet 2000, 356, 1371–1372, doi:10.1016/S0140-6736(00)02836-1.

17. Cohen, J.; E. Wakefield, C.; G. Laing, D. Smell and Taste Disorders Resulting from Cancer and Chemotherapy. Curr. Pharm. Des. 2016, 22, 2253–2263, doi:10.2174/1381612822666160216150812.

18. Nolden, A.; Joseph, P. V.; Kober, K.M.; Cooper, B.A.; Paul, S.M.; Hammer, M.J.; Dunn, L.B.; Conley, Y.P.; Levine, J.D.; Miaskowski, C. Co-Occurring Gastrointestinal Symptoms Are Associated With Taste Changes in Oncology Patients Receiving Chemotherapy. J. Pain Symptom Manage. 2019, 58, 756–765, doi:10.1016/j.jpainsymman.2019.07.016.

19. Özkan, İ.; Taylan, S.; Eroğlu, N.; Kolaç, N. The Relationship between Malnutrition and Subjective Taste Change Experienced by Patients with Cancer Receiving Outpatient Chemotherapy Treatment. Nutr. Cancer 2022, 74, 1670–1679, doi:10.1080/01635581.2021.1957485.

20. Bromley, S.M. Smell and Taste Disorders: A Primary Care Approach. Am Farm Physician 2000, 61, 427–436.

21. De Melo Silva, F.R.; De Oliveira, M.G.O.A.; Souza, A.S.R.; Figueroa, J.N.; Santos, C.S. Factors Associated with Malnutrition in Hospitalized Cancer Patients: A Croos-Sectional Study. Nutr. J. 2015, 14, 1–8, doi:10.1186/S12937-015-0113-1/TABLES/4.

22. Brown, D.; Loeliger, J.; Stewart, J.; Graham, K.L.; Goradia, S.; Gerges, C.; Lyons, S.; Connor, M.; Stewart, S.; Di Giovanni, A.;, et al. Relationship between Global Leadership Initiative on Malnutrition (GLIM) Defined Malnutrition and Survival, Length of Stay and Post-Operative Complications in People with Cancer: A Systematic Review. Clin. Nutr. 2023, 42, 255–268, doi:10.1016/j.clnu.2023.01.012.

23. Matsui, R.; Rifu, K.; Watanabe, J.; Inaki, N.; Fukunaga, T. Impact of Malnutrition as Defined by the GLIM Criteria on Treatment Outcomes in Patients with Cancer: A Systematic Review and Meta-Analysis. Clin. Nutr. 2023, 42, 615–624, doi:10.1016/J.CLNU.2023.02.019/ATTACHMENT/F4B5A529-8AB6-427A-A536-52258C2BBB25/MMC2.DOCX.

24. Xu, J.; Jie, Y.; Sun, Y.; Gong, D.; Fan, Y. Association of Global Leadership Initiative on Malnutrition with Survival Outcomes in Patients with Cancer: A Systematic Review and Meta-Analysis. Clin. Nutr. 2022, 41, 1874–1880, doi:10.1016/J.CLNU.2022.07.007.

25. de Vries, Y.C.; Winkels, R.M.; van den Berg, M.M.G.A.; de Graaf, C.; Kelfkens, C.S.; de Kruif, J.T.C.M.; Göker, E.; Grosfeld, S.; Sommeijer, D.W.; van Laarhoven, H.W.M.;, et al. Altered Food Preferences and Chemosensory Perception during Chemotherapy in Breast Cancer Patients: A Longitudinal Comparison with Healthy Controls. Food Qual. Prefer. 2018, 63, 135– 143, doi:10.1016/J.FOODQUAL.2017.09.003.

26. Gamper, E.M.; Giesinger, J.M.; Oberguggenberger, A.; Kemmler, G.; Wintner, L.M.; Gattringer, K.; Sperner-Unterweger, B.; Holzner, B.; Zabernigg, A. Taste Alterations in Breast and Gynaecological Cancer Patients Receiving Chemotherapy: Prevalence, Course of Severity, and Quality of Life Correlates. Acta Oncol. 2012, 51, 490–496, doi:10.3109/0284186X.2011.633554.

27. Berteretche, M. V.; Dalix, A.M.; D’Ornano, A.M.C.; Bellisle, F.; Khayat, D.; Faurion, A. Decreased Taste Sensitivity in Cancer Patients under Chemotherapy. Support. Care Cancer 2004, 12, 571–576, doi:10.1007/S00520-004-0589-2.

28. Ejder, Z.B.; Sanlier, N. The Relationship between Loneliness, Psychological Resilience, Quality of Life and Taste Change in Cancer Patients Receiving Chemotherapy. Support. Care Cancer 2023, 31, doi:10.1007/S00520-023-08156-W.

29. Comeau, T.B.; Epstein, J.B.; Migas, C. Taste and Smell Dysfunction in Patients Receiving Chemotherapy: A Review of Current Knowledge. Support. Care Cancer 2001, 9, 575–580, doi:10.1007/S005200100279/METRICS.

30. Ito, K.; Yuki, S.; Nakatsumi, H.; Kawamoto, Y.; Harada, K.; Nakano, S.; Saito, R.; Ando, T.; Sawada, K.; Yagisawa, M.;, et al. Multicenter, Prospective, Observational Study of Chemotherapy-Induced Dysgeusia in Gastrointestinal Cancer. Support. Care Cancer 2022, 30, 5351–5359, doi:10.1007/S00520-022-06936-4/TABLES/2.

31. Fujii, H.; Hirose, C.; Ishihara, M.; Iihara, H.; Imai, H.; Tanaka, Y.; Matsuhashi, N.; Takahashi, T.; Yamaguchi, K.; Yoshida, K.;, et al. Improvement of Dysgeusia by Polaprezinc, a Zinc-L-Carnosine, in Outpatients Receiving Cancer Chemotherapy. Anticancer Res. 2018, 38, 6367–6373, doi:10.21873/ANTICANRES.12995.

32. Strasser, F.; Demmer, R.; Böhme, C.; Schmitz, S.-F.H.; Thuerlimann, B.; Cerny, T.; Gillessen, S. Prevention of Docetaxel- or Paclitaxel-Associated Taste Alterations in Cancer Patients with Oral Glutamine: A Randomized, Placebo-Controlled, Double-Blind Study. Oncologist 2008, 13, 337–346, doi:10.1634/THEONCOLOGIST.2007-0217.

33. Turcotte, K.; Touchette, C.J.; Iorio-Morin, C.; Fortin, D. Successful Management of Glioblastoma Chemotherapy-Associated Dysgeusia with Gabapentin. Can. J. Neurol. Sci. 2020, 47, 839–841, doi:10.1017/CJN.2020.115.

34. Sevryugin, O.; Kasvis, P.; Vigano, M.L.; Vigano, A. Taste and Smell Disturbances in Cancer Patients: A Scoping Review of Available Treatments. Support. Care Cancer 2021, 29, 49–66, doi:10.1007/S00520-020-05609-4/TABLES/3.

35. Kurihara, K.; Beidler, L.M. Taste-Modifying Protein from Miracle Fruit. Science 1968, 161, 1241–1243, doi:10.1126/SCIENCE.161.3847.1241.

36. Misaka, T. Molecular Mechanisms of the Action of Miraculin, a Taste-Modifying Protein. Semin. Cell Dev. Biol. 2013, 24, 222–225, doi:10.1016/J.SEMCDB.2013.02.008.

37. Rodrigues, J.F.; Andrade, R. da S.; Bastos, S.C.; Coelho, S.B.; Pinheiro, A.C.M. Miracle Fruit: An Alternative Sugar Substitute in Sour Beverages. Appetite 2016, 107, 645–653, doi:10.1016/J.APPET.2016.09.014.

38. Penna, S. Chemotherapy-Induced Taste Alteration. Clin. J. Oncol. Nurs. 2023, 27, 479–485, doi:10.1188/23.CJON.479-485.

39. Soares, H.P.; Cusnir, M.; Schwartz, M.A.; Pizzolato, J.F.; Lutzky, J.; Campbell, R.J.; Beaumont, J.L.; Eton, D.; Stonick, S.; Lilenbaum, R. Treatment of Taste Alterations in Chemotherapy Patients Using the “Miracle Fruit”: Preliminary Analysis of a Pilot Study. https://doi.org/10.1200/jco.2010.28.15_suppl.e19523 2010, 28, e19523–e19523, doi:10.1200/JCO.2010.28.15_SUPPL.E19523.

40. Wilken, M.K.; Satiroff, B.A. Pilot Study of “Miracle Fruit” to Improve Food Palatability for Patients Receiving Chemotherapy. Clin. J. Oncol. Nurs. 2012, 16, doi:10.1188/12.CJON.E173-E177.

41. Turck, D.; Castenmiller, J.; De Henauw, S.; Hirsch-Ernst, K.I.; Kearney, J.; Maciuk, A.; Mangelsdorf, I.; McArdle, H.J.; Naska, A.; Pelaez, C.;, et al. Safety of Dried Fruits of Synsepalum Dulcificum as a Novel Food Pursuant to Regulation (EU) 2015/2283. EFSA journal. Eur. Food Saf. Auth. 2021, 19, doi:10.2903/J.EFSA.2021.6600.

42. López-Plaza, B.; Gil, Á.; Menéndez-Rey, A.; Bensadon-Naeder, L.; Hummel, T.; Feliú-Batlle, J.; Palma-Milla, S. Effect of Regular Consumption of a Miraculin-Based Food Supplement on Taste Perception and Nutritional Status in Malnourished Cancer Patients: A Triple-Blind, Randomized, Placebo-Controlled Clinical Trial-CLINMIR Pilot Protocol. Nutrients 2023, 15, doi:10.3390/NU15214639/S1.

43. Cederholm, T.; Jensen, G.L.; Correia, M.I.T.D.; Gonzalez, M.C.; Fukushima, R.; Higashiguchi, T.; Baptista, G.; Barazzoni, R.; Blaauw, R.; Coats, A.;, et al. GLIM Criteria for the Diagnosis of Malnutrition – A Consensus Report from the Global Clinical Nutrition Community. Clin. Nutr. 2019, 38, 1–9, doi:10.1016/j.clnu.2018.08.002.

44. Arbeitsgemeinschaft OuG, S. AWMF: Aktuelle Leitlinien 1996,.

45. Arraras, J.I.; Arias, F.; Tejedor, M.; Pruja, E.; Marcos, M.; Martínez, E.; Valerdi, J. The EORTC QLQ-C30 (Version 3.0) Quality of Life Questionnaire: Validation Study for Spain with Head and Neck Cancer Patients. Psychooncology. 2002, 11, 249–256, doi:10.1002/PON.555.

46. Cancer Institute, N. Common Terminology Criteria for Adverse Events (CTCAE) Common Terminology Criteria for Adverse Events (CTCAE) v5.0. 2017.

47. de la Torre-Aguilar, M.J.; Gomez-Fernandez, A.; Flores-Rojas, K.; Martin-Borreguero, P.; Mesa, M.D.; Perez-Navero, J.L.; Olivares, M.; Gil, A.; Gil-Campos, M. Docosahexaenoic and Eicosapentaenoic Intervention Modifies Plasma and Erythrocyte Omega-3 Fatty Acid Profiles But Not the Clinical Course of Children With Autism Spectrum Disorder: A Randomized Control Trial. Front. Nutr. 2022, 9, doi:10.3389/FNUT.2022.790250.

48. Lepage, G.; Roy, C.C. Specific Methylation of Plasma Nonesterified Fatty Acids in a One-Step Reaction. J. Lipid Res. 1988, 29, 227–235, doi:10.1016/S0022-2275(20)38553-9.

49. Hutton, J.L.; Baracos, V.E.; Wismer, W. V. Chemosensory Dysfunction Is a Primary Factor in the Evolution of Declining Nutritional Status and Quality of Life in Patients with Advanced Cancer. J. Pain Symptom Manage. 2007, 33, 156–165, doi:10.1016/J.JPAINSYMMAN.2006.07.017.

50. Brisbois, T.D.; Hutton, J.L.; Baracos, V.E.; Wismer, W.V. Taste and Smell Abnormalities as an Independent Cause of Failure of Food Intake in Patients with Advanced Cancer--an Argument for the Application of Sensory Science. J. Palliat. Care 2006, 22, 111–114, doi:10.1177/082585970602200208.

51. Obayashi, N.; Sugita, M.; Shintani, T.; Nishi, H.; Ando, T.; Kajiya, M.; Kawaguchi, H.; Ohge, H.; Naito, M. Taste-Taste Associations in Chemotherapy-Induced Subjective Taste Alterations: Findings from a Questionnaire Survey in an Outpatient Clinic. Support. Care Cancer 2023, 31, doi:10.1007/S00520-023-08013-W.

52. Pedersini, R.; Zamparini, M.; Bosio, S.; di Mauro, P.; Turla, A.; Monteverdi, S.; Zanini, A.; Amoroso, V.; Vassalli, L.; Cosentini, D.;, et al. Taste Alterations during Neo/Adjuvant Chemotherapy and Subsequent Follow-up in Breast Cancer Patients: A Prospective Single-Center Clinical Study. Support. Care Cancer 2022, 30, 6955–6961, doi:10.1007/S00520-022-07091-6.

53. Rehwaldt, M.; Wickham, R.; Purl, S.; Tariman, J.; Blendowski, C.; Shott, S.; Lappe, M. Self-Care Strategies to Cope with Taste Changes after Chemotherapy. Oncol. Nurs. Forum 2009, 36, doi:10.1188/09.ONF.E47-E56.

54. Ishikawa, T.; Morita, J.; Kawachi, K.; Tagashira, H. [Incidence of Dysgeusia Associated with Chemotherapy for Cancer]. Gan To Kagaku Ryoho. 2013, 40, 1049–1054.

55. Haemmerle, R.J.; Jatoi, A. Loss of Appetite in Patients with Cancer: An Update on Characterization, Mechanisms, and Palliative Therapeutics. Curr. Opin. Support. Palliat. Care 2023, 17, 168–171, doi:10.1097/SPC.0000000000000669.

56. de Vries, Y.C.; van den Berg, M.M.G.A.; de Vries, J.H.M.; Boesveldt, S.; de Kruif, J.T.C.M.; Buist, N.; Haringhuizen, A.; Los, M.; Sommeijer, D.W.; Timmer-Bonte, J.H.N.;, et al. Differences in Dietary Intake during Chemotherapy in Breast Cancer Patients Compared to Women without Cancer. Support. Care Cancer 2017, 25, 2581–2591, doi:10.1007/S00520-017-3668-X.

57. Yang, J.; Wei, H.; Zhou, Y.; Szeto, C.H.; Li, C.; Lin, Y.; Coker, O.O.; Lau, H.C.H.; Chan, A.W.H.; Sung, J.J.Y.;, et al. High-Fat Diet Promotes Colorectal Tumorigenesis Through Modulating Gut Microbiota and Metabolites. Gastroenterology 2022, 162, 135–149.e2, doi:10.1053/J.GASTRO.2021.08.041.

58. Tong, Y.; Gao, H.; Qi, Q.; Liu, X.; Li, J.; Gao, J.; Li, P.; Wang, Y.; Du, L.; Wang, C. High Fat Diet, Gut Microbiome and Gastrointestinal Cancer. Theranostics 2021, 11, 5889–5910, doi:10.7150/THNO.56157.

59. Shao, X.; Liu, L.; Zhou, Y.; Zhong, K.; Gu, J.; Hu, T.; Yao, Y.; Zhou, C.; Chen, W. High-Fat Diet Promotes Colitis-Associated Tumorigenesis by Altering Gut Microbial Butyrate Metabolism. Int. J. Biol. Sci. 2023, 19, 5004–5019, doi:10.7150/IJBS.86717.

60. Hayashi, T.; Fujita, K.; Nojima, S.; Hayashi, Y.; Nakano, K.; Ishizuya, Y.; Wang, C.; Yamamoto, Y.; Kinouchi, T.; Matsuzaki, K.;, et al. High-Fat Diet-Induced Inflammation Accelerates Prostate Cancer Growth via IL6 Signaling. Clin. Cancer Res. 2018, 24, 4309–4318, doi:10.1158/1078-0432.CCR-18-0106.

61. Zhang, J.; Guo, S.; Li, J.; Bao, W.; Zhang, P.; Huang, Y.; Ling, P.; Wang, Y.; Zhao, Q. Effects of High-Fat Diet-Induced Adipokines and Cytokines on Colorectal Cancer Development. FEBS Open Bio 2019, 9, 2117–2125, doi:10.1002/2211-5463.12751.

62. Arita, S.; Kinoshita, Y.; Ushida, K.; Enomoto, A.; Inagaki-Ohara, K. High-Fat Diet Feeding Promotes Stemness and Precancerous Changes in Murine Gastric Mucosa Mediated by Leptin Receptor Signaling Pathway. Arch. Biochem. Biophys. 2016, 610, 16–24, doi:10.1016/J.ABB.2016.09.015.

63. Ruan, L.; Cheng, S.P.; Zhu, Q.X. Dietary Fat Intake and the Risk of Skin Cancer: A Systematic Review and Meta-Analysis of Observational Studies. Nutr. Cancer 2020, 72, 398–408, doi:10.1080/01635581.2019.1637910.

64. Kim, Y.; Je, Y.; Giovannucci, E.L. Association between Dietary Fat Intake and Mortality from All-Causes, Cardiovascular Disease, and Cancer: A Systematic Review and Meta-Analysis of Prospective Cohort Studies. Clin. Nutr. 2021, 40, 1060–1070, doi:10.1016/J.CLNU.2020.07.007.

65. Hunter, D.J.; Spiegelman, D.; Adami, H.-O.; Beeson, L.; van den Brandt, P.A.; Folsom, A.R.; Fraser, G.E.; Goldbohm, R.A.; Graham, S.; Howe, G.R.;, et al. Cohort Studies of Fat Intake and the Risk of Breast Cancer--a Pooled Analysis. N. Engl. J. Med. 1996, 334, 356–361, doi:10.1056/NEJM199602083340603.

66. Cao, D. Xing; Wu, G. Hao; Zhang, B.; Quan, Y. Jun; Wei, J.; Jin, H.; Jiang, Y.; Yang, Z. ang Resting Energy Expenditure and Body Composition in Patients with Newly Detected Cancer. Clin. Nutr. 2010, 29, 72–77, doi:10.1016/J.CLNU.2009.07.001.

67. Brennan, S.F.; Woodside, J. V.; Lunny, P.M.; Cardwell, C.R.; Cantwell, M.M. Dietary Fat and Breast Cancer Mortality: A Systematic Review and Meta-Analysis. Crit. Rev. Food Sci. Nutr. 2017, 57, 1999–2008, doi:10.1080/10408398.2012.724481.

68. Bojková, B.; Winklewski, P.J.; Wszedybyl-Winklewska, M. Dietary Fat and Cancer-Which Is Good, Which Is Bad, and the Body of Evidence. Int. J. Mol. Sci. 2020, 21, 1–56, doi:10.3390/IJMS21114114.

69. Sellem, L.; Srour, B.; Guéraud, F.; Pierre, F.; Kesse-Guyot, E.; Fiolet, T.; Lavalette, C.; Egnell, M.; Latino-Martel, P.; Fassier, P.;, et al. Saturated, Mono- and Polyunsaturated Fatty Acid Intake and Cancer Risk: Results from the French Prospective Cohort NutriNet-Santé. Eur. J. Nutr. 2019, 58, 1515–1527, doi:10.1007/S00394-018-1682-5.

70. Guasch-Ferré, M.; Zong, G.; Willett, W.C.; Zock, P.L.; Wanders, A.J.; Hu, F.B.; Sun, Q. Associations of Monounsaturated Fatty Acids From Plant and Animal Sources With Total and Cause-Specific Mortality in Two US Prospective Cohort Studies. Circ. Res. 2019, 124, 1266–1275, doi:10.1161/CIRCRESAHA.118.313996.

71. Gunstone, F.D. Fatty Acids—Nomenclature, Structure, Isolation and Structure Deter-Mination, Biosynthesis and Chemical Synthesis.Tle. In Fatty Acid and Lipid Chemistry; Springer US: Boston, MA, USA., 1996; pp. 1–34.

72. Psaltopoulou, T.; Kosti, R.I.; Haidopoulos, D.; Dimopoulos, M.; Panagiotakos, D.B. Olive Oil Intake Is Inversely Related to Cancer Prevalence: A Systematic Review and a Meta-Analysis of 13,800 Patients and 23,340 Controls in 19 Observational Studies. Lipids Health Dis. 2011, 10, doi:10.1186/1476-511X-10-127.

73. Yao, X.; Tian, Z. Saturated, Monounsaturated and Polyunsaturated Fatty Acids Intake and Risk of Pancreatic Cancer: Evidence from Observational Studies. PLoS One 2015, 10, doi:10.1371/JOURNAL.PONE.0130870.

74. Wu, Q.J.; Gong, T.T.; Wang, Y.Z. Dietary Fatty Acids Intake and Endometrial Cancer Risk: A Dose-Response Meta-Analysis of Epidemiological Studies. Oncotarget 2015, 6, 36081–36097, doi:10.18632/ONCOTARGET.5555.

75. Harris, W.S.; Von Schacky, C. The Omega-3 Index: A New Risk Factor for Death from Coronary Heart Disease? Prev. Med. (Baltim). 2004, 39, 212–220, doi:10.1016/j.ypmed.2004.02.030.

76. Chen, Q.; Wang, J.; Wang, J.; Lin, J.; Chen, L.; Lin, L. Song; Pan, L. zhen; Shi, B.; Qiu, Y.; Zheng, X. yan; et al. Erythrocyte ω-3 Polyunsaturated Fatty Acids Are Inversely Associated with the Risk of Oral Cancer: A Case-Control Study. Nutr. Diabetes 2020, 10, doi:10.1038/S41387-020-00140-1.

77. Hanson, S.; Thorpe, G.; Winstanley, L.; Abdelhamid, A.S.; Hooper, L.; Abdelhamid, A.; Ajabnoor, S.; Alabdulghafoor, F.; Alkhudairy, L.; Biswas, P.;, et al. Omega-3, Omega-6 and Total Dietary Polyunsaturated Fat on Cancer Incidence: Systematic Review and Meta-Analysis of Randomised Trials. Br. J. Cancer 2020, 122, 1260–1270, doi:10.1038/S41416-020-0761-6.

78. Koundouros, N.; Poulogiannis, G. Reprogramming of Fatty Acid Metabolism in Cancer. Br. J. Cancer 2020, 122, 4–22, doi:10.1038/S41416-019-0650-Z.

79. Amézaga, J.; Arranz, S.; Urruticoechea, A.; Ugartemendia, G.; Larraioz, A.; Louka, M.; Uriarte, M.; Ferreri, C.; Tueros, I. Altered Red Blood Cell Membrane Fatty Acid Profile in Cancer Patients. Nutrients 2018, 10, doi:10.3390/NU10121853.

80. Hamaker, M.E.; Oosterlaan, F.; van Huis, L.H.; Thielen, N.; Vondeling, A.; van den Bos, F. Nutritional Status and Interventions for Patients with Cancer - A Systematic Review. J. Geriatr. Oncol. 2021, 12, 6–21, doi:10.1016/J.JGO.2020.06.020.

81. Fuentes, N.R.; Kim, E.; Fan, Y.Y.; Chapkin, R.S. Omega-3 Fatty Acids, Membrane Remodeling and Cancer Prevention. Mol. Aspects Med. 2018, 64, 79–91, doi:10.1016/J.MAM.2018.04.001.

82. Serhan, C.N.; Sulciner, M.L. Resolution Medicine in Cancer, Infection, Pain and Inflammation: Are We on Track to Address the next Pandemic? Cancer Metastasis Rev. 2023, 42, 13–17, doi:10.1007/S10555-023-10091-5.

83. Dyall, S.C.; Balas, L.; Bazan, N.G.; Brenna, J.T.; Chiang, N.; da Costa Souza, F.; Dalli, J.; Durand, T.; Galano, J.M.; Lein, P.J.;, et al. Polyunsaturated Fatty Acids and Fatty Acid-Derived Lipid Mediators: Recent Advances in the Understanding of Their Biosynthesis, Structures, and Functions. Prog. Lipid Res. 2022, 86, doi:10.1016/J.PLIPRES.2022.101165.

84. Serhan, C.N.; Chiang, N.; Dalli, J. The Resolution Code of Acute Inflammation: Novel pro-Resolving Lipid Mediators in Resolution. Semin. Immunol. 2015, 27, 200–215, doi:10.1016/J.SMIM.2015.03.004.

85. Freitas, R.D.S.; Campos, M.M. Protective Effects of Omega-3 Fatty Acids in Cancer-Related Complications. Nutrients 2019, 11, doi:10.3390/NU11050945.

86. Körner, A.; Schlegel, M.; Theurer, J.; Frohnmeyer, H.; Adolph, M.; Heijink, M.; Giera, M.; Rosenberger, P.; Mirakaj, V. Resolution of Inflammation and Sepsis Survival Are Improved by Dietary Ω-3 Fatty Acids. Cell Death Differ. 2018, 25, 421– 431, doi:10.1038/CDD.2017.177.

87. Saini, R.K.; Prasad, P.; Sreedhar, R.V.; Naidu, K.A.; Shang, X.; Keum, Y.S. Omega-3 Polyunsaturated Fatty Acids (PUFAs): Emerging Plant and Microbial Sources, Oxidative Stability, Bioavailability, and Health Benefits-A Review. *Antioxidants (Basel*, Switzerland) 2021, 10, doi:10.3390/ANTIOX10101627.

88. Thiagarajan, P.; Parker, C.J.; Prchal, J.T. How Do Red Blood Cells Die? Front. Physiol. 2021, 12, doi:10.3389/FPHYS.2021.655393.

89. Breitkreutz, R.; Tesdal, K.; Jentschura, D.; Haas, O.; Leweling, H.; Holm, E. Effects of a High-Fat Diet on Body Composition in Cancer Patients Receiving Chemotherapy: A Randomized Controlled Study. Wien. Klin. Wochenschr. 2005, 117, 685–692, doi:10.1007/S00508-005-0455-3.

90. Kyle, U.G.; Bosaeus, I.; De Lorenzo, A.D.; Deurenberg, P.; Elia, M.; Gómez, J.M.; Heitmann, B.L.; Kent-Smith, L.; Melchior, J.C.; Pirlich, M.;, et al. Bioelectrical Impedance Analysis-Part II: Utilization in Clinical Practice. Clin. Nutr. 2004, 23, 1430–1453, doi:10.1016/J.CLNU.2004.09.012.

91. Bozzetti, F.; Pagnoni, A.M.; Del Vecchio, M. Excessive Caloric Expenditure as a Cause of Malnutrition in Patients with Cancer. Surg. Gynecol. Obstet. 1980, 150, 229–234.

92. Lim, H.S.; Lee, B.; Cho, I.; Cho, G.S. Nutritional and Clinical Factors Affecting Weight and Fat-Free Mass Loss after Gastrectomy in Patients with Gastric Cancer. Nutrients 2020, 12, 1–13, doi:10.3390/NU12071905.

93. Baracos, V.; Kazemi-Bajestani, S.M.R. Clinical Outcomes Related to Muscle Mass in Humans with Cancer and Catabolic Illnesses. Int. J. Biochem. Cell Biol. 2013, 45, 2302–2308, doi:10.1016/J.BIOCEL.2013.06.016.

94. Martin, L.; Birdsell, L.; MacDonald, N.; Reiman, T.; Clandinin, M.T.; McCargar, L.J.; Murphy, R.; Ghosh, S.; Sawyer, M.B.; Baracos, V.E. Cancer Cachexia in the Age of Obesity: Skeletal Muscle Depletion Is a Powerful Prognostic Factor, Independent of Body Mass Index. J. Clin. Oncol. 2013, 31, 1539–1547, doi:10.1200/JCO.2012.45.2722.

95. Sadeghi, M.; Keshavarz-Fathi, M.; Baracos, V.; Arends, J.; Mahmoudi, M.; Rezaei, N. Cancer Cachexia: Diagnosis, Assessment, and Treatment. Crit. Rev. Oncol. Hematol. 2018, 127, 91–104, doi:10.1016/J.CRITREVONC.2018.05.006.

96. Lewandowska, A.; Rudzki, G.; Lewandowski, T.; Próchnicki, M.; Rudzki, S.; Laskowska, B.; Brudniak, J. Quality of Life of Cancer Patients Treated with Chemotherapy. Int. J. Environ. Res. Public Health 2020, 17, 1–16, doi:10.3390/IJERPH17196938.

97. Polański, J.; Chabowski, M.; Świątoniowska-Lonc, N.; Dudek, K.; Jankowska-Polańska, B.; Zabierowski, J.; Mazur, G. Relationship between Nutritional Status and Clinical Outcome in Patients Treated for Lung Cancer. Nutrients 2021, 13, doi:10.3390/NU13103332.

98. Maia, F. de C.P.; Silva, T.A.; Generoso, S. de V.; Correia, M.I.T.D. Malnutrition Is Associated with Poor Health-Related Quality of Life in Surgical Patients with Gastrointestinal Cancer. Nutrition 2020, 75–76, doi:10.1016/J.NUT.2020.110769.

99. Firkins, J.; Hansen, L.; Driessnack, M.; Dieckmann, N. Quality of Life in “Chronic” Cancer Survivors: A Meta-Analysis. J. Cancer Surviv. 2020, 14, 504–517, doi:10.1007/S11764-020-00869-9.

100. Fearon, K.; Strasser, F.; Anker, S.D.; Bosaeus, I.; Bruera, E.; Fainsinger, R.L.; Jatoi, A.; Loprinzi, C.; MacDonald, N.; Mantovani, G.;, et al. Definition and Classification of Cancer Cachexia: An International Consensus. Lancet. Oncol. 2011, 12, 489–495, doi:10.1016/S1470-2045(10)70218-7.

101. Salvetti, M. de G.; Machado, C.S.P.; Donato, S.C.T.; da Silva, A.M. Prevalence of Symptoms and Quality of Life of Cancer Patients. Rev. Bras. Enferm. 2020, 73, doi:10.1590/0034-7167-2018-0287.

102. Juhé-Beaulaton, D. Le Fruit Miracle (Synsepalum Dulcificum): Des Voyageurs Sur La Cote Ouest Africaine Auxlaboratoires Pharmaceutiques.; Paris, France, 2014; Vol. 11;.

103. Wong, J.M.; Kern, M. Miracle Fruit Improves Sweetness of a Low-Calorie Dessert without Promoting Subsequent Energy Compensation. Appetite 2011, 56, 163–166, doi:10.1016/J.APPET.2010.10.005.

104. Capitanio, A.; Lucci, G.; Tommasi, L. MIXING TASTE ILLUSIONS: THE EFFECT OF MIRACULIN ON BINARY AND TRINARY MIXTURES. J. Sens. Stud. 2011, 26, 54–61, doi:10.1111/J.1745-459X.2010.00321.X.

105. Igarashi, G.; Higuchi, R.; Yamazaki, T.; Ito, N.; Ashida, I.; Miyaoka, Y. Differential Sweetness of Commercial Sour Liquids Elicited by Miracle Fruit in Healthy Young Adults. http://dx.doi.org/10.1177/1082013212443060 2013, 19, 243–249, doi:10.1177/1082013212443060.

106. Hudson, S.D.; Sims, C.A.; Odabasi, A.Z.; Colquhoun, T.A.; Snyder, D.J.; Stamps, J.J.; Dotson, S.C.; Puentes, L.; Bartoshuk, L.M. Flavor Alterations Associated with Miracle Fruit and Gymnema Sylvestre. Chem. Senses 2018, 43, 481–488, doi:10.1093/CHEMSE/BJY032.

107. Andrade, A.C.; Martins, M.B.; Rodrigues, J.F.; Coelho, S.B.; Pinheiro, A.C.M.; Bastos, S.C. Effect of Different Quantities of Miracle Fruit on Sour and Bitter Beverages. LWT 2019, 99, 89–97, doi:10.1016/J.LWT.2018.09.054.

108. Tafazoli, S.; Vo, T.D.; Roberts, A.; Rodriguez, C.; Viñas, R.; Madonna, M.E.; Chiang, Y.H.; Noronha, J.W.; Holguin, J.C.; Ryder, J.A.;, et al. Safety Assessment of Miraculin Using in Silico and in Vitro Digestibility Analyses. Food Chem. Toxicol. 2019, 133, doi:10.1016/J.FCT.2019.110762.

109. Choi, S.E.; Garza, J. Effects of Different Miracle Fruit Products on the Sensory Characteristics of Different Types of Sour Foods by Descriptive Analysis. J. Food Sci. 2020, 85, 36–49, doi:10.1111/1750-3841.14988.

110. Temussi, P.A. Natural Sweet Macromolecules: How Sweet Proteins Work. Cell. Mol. Life Sci. 2006, 63, 1876–1888, doi:10.1007/S00018-006-6077-8.

111. Kurihara, Y. Characteristics of Antisweet Substances, Sweet Proteins, and Sweetness-Inducing Proteins. Crit. Rev. Food Sci. Nutr. 1992, 32, 231–252, doi:10.1080/10408399209527598.

112. Huang, W.; Chung, H.Y.; Xuan, W.; Wang, G.; Li, Y. The Cholesterol-Lowering Activity of Miracle Fruit (Synsepalum Dulcificum). J. Food Biochem. 2020, 44, e13185, doi:10.1111/JFBC.13185.

113. Haddad, S.G.; Mohammad, M.; Raafat, K.; Saleh, F.A. Antihyperglycemic and Hepatoprotective Properties of Miracle Fruit (Synsepalum Dulcificum) Compared to Aspartame in Alloxan-Induced Diabetic Mice. J. Integr. Med. 2020, 18, 514–521, doi:10.1016/J.JOIM.2020.09.001.

114. Obafemi, T.O.; Olaleye, M.T.; Akinmoladun, A.C. Antidiabetic Property of Miracle Fruit Plant (Synsepalum Dulcificum Shumach. & Thonn. Daniell) Leaf Extracts in Fructose-Fed Streptozotocin-Injected Rats via Anti-Inflammatory Activity and Inhibition of Carbohydrate Metabolizing Enzymes. J. Ethnopharmacol. 2019, 244, 112124–112124, doi:10.1016/J.JEP.2019.112124.

115. Chen, C.C.; Liu, I.M.; Cheng, J.T. Improvement of Insulin Resistance by Miracle Fruit (Synsepalum Dulcificum) in Fructose-Rich Chow-Fed Rats. Phyther. Res. 2006, 20, 987–992, doi:10.1002/PTR.1919.

116. Shi, Y.C.; Lin, K.S.; Jhai, Y.F.; Lee, B.H.; Han, Y.; Cui, Z.; Hsu, W.H.; Wu, S.C. Miracle Fruit (Synsepalum Dulcificum) Exhibits as a Novel Anti-Hyperuricaemia Agent. Molecules 2016, 21, doi:10.3390/MOLECULES21020140.

