## Supplemental Table S1-S4 for "Efficacy and Safety of Habitual Consumption of a Food Supplement Containing Miraculin in Malnourished Cancer Patients: the CLINMIR Pilot Study"

*Article (Supplementary Materials)*

**Table S1.** Perception of food consumption depending on treatment

| | | Standard dose of DMB | | | High dose of DMB | | | Placebo | | | P-value | | |
|---|---|---|---|---|---|---|---|---|---|---|---|---|---|
| | | 1 month | 2 months | 3 months | 1 month | 2 months | 3 months | 1 month | 2 months | 3 months | 1 month | 2 months | 3 months |
| Eat the same | Yes (%) | 66.7 | 85.7 | 50 | 85.7 | 50 | 25 | 50 | 50 | 50 | 0.348 | 0.298 | 0.683 |
| | No (%) | 33.3 | 14.3 | 50 | 14.3 | 50 | 75 | 50 | 50 | 50 | | | |
| Eat less | Yes (%) | 22.2 | 0 | 0 | 0 | 33.3 | 50 | 0 | 0 | 0 | 0.204 | 0.089 | 0.032 |
| | No (%) | 77.8 | 100 | 100,00 | 100 | 66.7 | 50 | 100 | 100 | 100 | | | |
| Eat more | Yes (%) | 11.1 | 14.3 | 50,00 | 14.3 | 16.7 | 25 | 50 | 50 | 50 | 0.172 | 0.280 | 0.683 |
| | No (%) | 88.9 | 85.7 | 50,00 | 85.7 | 83.3 | 75 | 50 | 50 | 50 | | | |

**Table S2.** Nutritional status depending on treatment (%)

| | | Standard dose of DMB | | High dose of DMB | | Placebo | | p-value | |
|---|---|---|---|---|---|---|---|---|---|
| | | Baseline | 3 months | Baseline | 3 months | Baseline | 3 months | Baseline | 3 months |
| Weight lost percentage | No | 0 | 87.5 | 0 | 83.3 | 0 | 100 | | |
| | 5 – 10 | 70 | 12.5 | 81.8 | 16.7 | 70 | 0 | 0.772 | 0.556 |
| | >10 | 30 | 0 | 18.2 | 0 | 30 | 0 | | |
| GLIM Criteria Malnutrition | Normal | 0 | 62.5 | 0 | 83.3 | 0 | 71.4 | | |
| | Moderate | 50 | 12.5 | 81.8 | 16.7 | 70 | 28.6 | 0.556 | 0.407 |
| | Severe | 50 | 25 | 18.2 | 0 | 30 | 0 | | |

**Table S3.** Perceived effectiveness of the product depending on treatment (mean ± SD)

| | | Standard dose of DMB | | | | High dose of DMB | | | | Placebo | | | | P-value | | |
|---|---|---|---|---|---|---|---|---|---|---|---|---|---|---|---|---|
| | | Baseline | 1 week | 2 months | 3 months | Baseline | 1 week | 2 meses | 3 meses | Baseline | 1 week | 2 meses | 3 meses | Time (t) | Treatment (T) | T x t |
| Perceived effectiveness | Points | 33.0 ± 27.5 | 33.0 ± 27.5 | 62.5 ± 23.8 | 63.8 ± 21.3 | 46.5 ± 25.5 | 44.5 ± 27.2 | 71.7 ± 24.2 | 58.3 ± 37.2 | 66.4 ± 42.9 | 59.3 ± 40.5 | 75.0 ± 38.9 | 75.0 ± 38.9 | 0.052 | 0.074 | 0.628 |

**Table S4.** Adverse events depending on the assigned treatment group (%)

| | | Standard dose of DMB | | | High dose of DMB | | | Placebo | | | P-value | | |
|---|---|---|---|---|---|---|---|---|---|---|---|---|---|
| | | 1 month | 2 months | 3 months | 1 month | 2 months | 3 months | 1 month | 2 months | 3 months | 1 month | 2 months | 3 months |
| Abdominal distension | Grade 0 | 90 | 87.5 | 100 | 85.7 | 66.7 | 60 | 100 | 83.3 | 83.3 | | | |
| | Grade 1 | 10 | 12.5 | 0 | 14.3 | 16.7 | 40 | 0 | 16.7 | 16.7 | 0.649 | 0.629 | 0.157 |
| | Grade 2 | 0 | 0 | 0 | 0 | 16.7 | 0 | 0 | 0 | 0 | | | |
| Abdominal pain | Grade 0 | 90 | 100 | 87.5 | 85.7 | 100 | 60 | 83.3 | 100 | 83.3 | | | |
| | Grade 1 | 10 | 0 | 12.5 | 14.3 | 0 | 20 | 16.7 | 0 | 16.7 | 0.578 | 1 | 0.520 |
| | Grade 2 | 0 | 0 | 0 | 0 | 0 | 20 | 0 | 0 | 0 | | | |
| Nausea | Grade 0 | 90 | 100 | 87.5 | 85.7 | 83.3 | 80 | 100 | 100 | 100 | | | |
| | Grade 1 | 10 | 0 | 12.5 | 14.3 | 16.7 | 20 | 0 | 0 | 0 | 0.454 | 0.293 | 0.545 |
| | Grade 2 | 0 | 0 | 0 | 0 | 0 | 0 | 0 | 0 | 0 | | | |
| Regurgitation | Grade 0 | 90 | 100 | 75 | 85.7 | 100 | 60 | 100 | 83.3 | 100 | | | |
| | Grade 1 | 10 | 0 | 25 | 14.3 | 0 | 20 | 0 | 16.7 | 0 | 0.454 | 0.293 | 0.307 |
| | Grade 2 | 0 | 0 | 0 | 0 | 0 | 20 | 0 | 0 | 0 | | | |
| Vomiting | Grade 0 | 100 | 100 | 100 | 85.7 | 83.3 | 80 | 83.3 | 100 | 100 | | | |
| | Grade 1 | 0 | 0 | 0 | 14.3 | 16.7 | 20 | 16.7 | 0 | 0 | 0.426 | 0.293 | 0.228 |
| | Grade 2 | 0 | 0 | 0 | 0 | 0 | 0 | 0 | 0 | 0 | | | |
| Constipation | Grade 0 | 90 | 100 | 87.5 | 83.3 | 66.7 | 40 | 100 | 100 | 66.7 | | | |
| | Grade 1 | 10 | 0 | 12.5 | 16.7 | 33.3 | 40 | 0 | 0 | 33.3 | 0.598 | 0.075 | 0.301 |
| | Grade 2 | 0 | 0 | 0 | 0 | 0 | 20 | 0 | 0 | 0 | | | |
| Diarrhea | Grade 0 | 100 | 100 | 100 | 57.1 | 100 | 60 | 100 | 100 | 83.3 | | | |
| | Grade 1 | 0 | 0 | 0 | 42.9 | 0 | 40 | 0 | 0 | 16.7 | 0.019 | 1 | 0.157 |
| | Grade 2 | 0 | 0 | 0 | 0 | 0 | 0 | 0 | 0 | 0 | | | |
| Flatulence | Grade 0 | 70 | 87.5 | 75 | 85.7 | 50 | 40 | 100 | 100 | 83.3 | | | |
| | Grade 1 | 30 | 12.5 | 25 | 14.3 | 33.3 | 40 | 0 | 0 | 16.7 | 0.158 | 0.227 | 0.378 |
| | Grade 2 | 0 | 0 | 0 | 0 | 16.7 | 20 | 0 | 0 | 0 | | | |
| Intensity | Grade 0 | 28.6 | 100 | 50 | 40 | 33.3 | 50 | 57.1 | 83.3 | 100 | | | |
| | Grade 1 | 14.3 | 0 | 50 | 20 | 0 | 50 | 42.9 | 16.7 | 0 | 0.346 | 0.096 | 0.247 |
| | Grade 2 | 57.1 | 0 | 0 | 40 | 66.7 | 0 | 0 | 0 | 0 | | | |
| Product relationship | Grade 0 | 33.3 | 100 | 100 | 100 | 100 | 100 | 85.7 | 100 | 100 | | | |
| | Grade 1 | 50 | 0 | 0 | 0 | 0 | 0 | 14.3 | 0 | 0 | 0.118 | 1 | 1 |
| | Grade 2 | 16.7 | 0 | 0 | 0 | 0 | 0 | 0 | 0 | 0 | | | |
| Conduct adopted | Grade 0 | 33.3 | 0 | 0 | 50 | 0 | 0 | 33.3 | 0 | 0 | | | |
| | Grade 1 | 50 | 100 | 100 | 50 | 0 | 100 | 66.7 | 100 | 100 | 0.833 | 0.135 | 1 |
| | Grade 2 | 16.7 | 0 | 0 | 0 | 100 | 0 | 0 | 0 | 0 | | | |

Grade 0, not described; Grade 1, mild; Grade 2, moderate.
